# Population Performance and Individual Agreement of Coronary Artery Disease Polygenic Risk Scores

**DOI:** 10.1101/2024.07.25.24310931

**Authors:** Sarah A. Abramowitz, Kristin Boulier, Karl Keat, Katie M. Cardone, Manu Shivakumar, John DePaolo, Renae Judy, Dokyoon Kim, Daniel J. Rader, Marylyn D. Ritchie, Benjamin F. Voight, Bogdan Pasaniuc, Michael G. Levin, Scott M. Damrauer

## Abstract

**Importance:** Polygenic risk scores (PRSs) for coronary artery disease (CAD) are a growing clinical and commercial reality. Whether existing scores provide similar individual-level assessments of disease liability is a critical consideration for clinical implementation that remains uncharacterized.

**Objective:** Characterize the reliability of CAD PRSs that perform equivalently at the population level at predicting individual-level risk.

**Design:** Cross-sectional Study.

**Setting:** All of Us Research Program (AOU), Penn Medicine Biobank (PMBB), and UCLA ATLAS Precision Health Biobank.

**Participants:** Volunteers of diverse genetic backgrounds enrolled in AOU, PMBB, and UCLA with available electronic health record and genotyping data.

**Exposures:** Polygenic risk for CAD from previously published PRSs and new PRSs developed separately from the testing cohorts.

**Main Outcomes and Measures:** Sets of CAD PRSs that perform population prediction equivalently were identified by comparing calibration and discrimination (Brier score and AUROC) of generalized linear models of prevalent CAD using Bayesian analysis of variance. Among equivalently performing scores, individual-level agreement between risk estimates was tested with intraclass correlation (ICC) and Light’s Kappa, measures of inter-rater reliability.

**Results:** 50 PRSs were calculated for 171,095 AOU participants. When included in a model of prevalent CAD, 48 scores had practically equivalent Brier scores and AUROCs (region of practical equivalence = 0.02). Across these scores, 84% of participants had at least one score in both the top and bottom risk quintile. Continuous agreement of individual risk predictions from the 48 scores was poor, with an ICC of 0.351 (95% CI; 0.349, 0.352). Agreement between two statistically equivalent scores was moderate, with an ICC of 0.649 (95% CI; 0.646, 0.652). Light’s Kappa, used to evaluate consistency of assignment to high-risk thresholds, did not exceed 0.56 (interpreted as ‘fair’) across statistically and practically equivalent scores. Repeating the analysis among 41,193 PMBB and 50,748 UCLA participants yielded different sets of statistically and practically equivalent scores which also lacked strong individual agreement.

**Conclusions and Relevance:** Across three diverse biobanks, CAD PRSs that performed equivalently at the population level produced unreliable individual risk estimates. Approaches to clinical implementation of CAD PRSs must consider the potential for discordant individual risk estimates from otherwise indistinguishable scores.

## Introduction

Polygenic risk scores (PRSs), which estimate an individual’s genetic liability for disease, have been proposed as a tool for improving prevention and treatment of coronary artery disease (CAD).^1^ This is grounded in an understanding that an estimated 40-60% of CAD susceptibility is attributable to genetics, and that ones’ genotype is fixed at conception.^2^ Although the genetic underpinnings of CAD are not yet comprehensively established, large-scale efforts continue to identify CAD-associated variation. Polygenic risk scores (PRS) aggregate the effects of many risk variants to predict the genetic component of one’s disease risk. Advocates propose that PRSs have the potential to enable early and precise identification of individuals at increased risk for CAD, and facilitate implementation of focused primary prevention as part of precision cardiovascular medicine initiatives.^3^ These applications include combining PRSs with clinical variables into a comprehensive risk models, considering a PRS as a “risk enhancer” to be applied to individuals at borderline risk, or considering a PRS risk estimate as a stand-alone test.^4^ Although use of CAD PRSs in any form is not currently the standard of care,^1^ it is nevertheless a growing reality, increasingly trialed by academic medical centers and major consortia,^5–7^ and offered clinically via physician-based and direct-to-consumer genetic testing companies.^4,8^

Advances in statistical techniques and the size and diversity of the genetic datasets used to construct PRSs continue to fuel the development of novel scores. To date, dozens of CAD PRS have been deposited in the PGS Catalog, which seeks to standardize and improve the reporting of PRSs. Proprietary commercial scores, for which the underlying genetic association data, methodology and weights are unknown and unregulated, are also being marketed.^9,10^ The growing menu of unique scores available for research, clinical, and commercial purposes presents patients and providers with the challenge of distinguishing between them.

Consensus recommendations from experts have been developed to describe the population-level evaluation of PRSs with consideration of performance measures of discrimination and calibration^11–13^ that can facilitate comparisons across PRSs in a given setting. These metrics provide a framework to assess how scores can effectively estimate risk at the population level, and how future scores can be compared to each other and improved over time. However, the degree to which different scores predict an individual’s underlying genetic liability to disease accurately and consistently is an important angle that has been largely overlooked. Whenever a clinical test is ordered for an individual – from blood panels to imaging – it is reasonable to expect that any qualified person or machine performing the test for that individual should produce similar results (with a tolerable level of error). In other words, these tests should be *reliable*. Reliability is a core consideration when evaluating the utility of clinical tests.^14^ Whether multiple PRSs for the same disease tend to provide similar individual-level assessments of disease liability remains largely uncharacterized. ^15^

To compare the reliability of risk assessment between PRSs, we sought to evaluate the population-level performance of available CAD PRSs, and among the set of equivalently performing scores, assess individual-level agreement of their risk estimates. We implemented this framework by analyzing risk predictions generated from new and existing PRSs across a total of 263,036 participants from three diverse biobanks.

## Methods

### Study Population

The primary analysis was conducted using data from the All of Us (AOU) Research Program, a National Institutes of Health-funded biobank composed of adult volunteers across the United States who have given written consent for analysis of their deidentified EHR and genetic data.^16,17^ At the time of analysis, 237,568 participants had available whole genome sequencing. For each participant, genetic sex was imputed using PLINK’s check_sex function. A binary CAD phenotype was assigned in the presence of at least one of the following codes: 410, 411, 412, 413, 41, V45.81 (ICD-9), I21, I22, I24, Z95.1, Z98.61, I20.0 (ICD-10). Age was calculated using participant birth year and the most recent data release cutoff date. Individuals without EHR data were excluded. All analyses were performed on deidentified data from All of Us Controlled Tier Dataset v7 by authorized researchers. Similar approaches were used for the Penn Medicine Biobank (PMBB) and UCLA ATLAS Precision Health Biobank (UCLA) (eMethods).^18–20^

### Selection and Creation of Polygenic Risk Scores

Polygenic risk scores for CAD were selected from the Polygenic Score (PGS) Catalog (eMethods).^9^ In addition, two novel CAD PRSs were created by applying Bayesian approaches – PRS-CSx-auto and LDPred2-auto – to summary statistics from a multi-population meta-analysis of genome-wide association studies (GWAS), comprising 380,508 individuals with and 1,836,455 individuals without CAD (eFigure 1); these scores were named “PGS_LDP2Auto” and “PGS_prscsx,” after the methods used to construct them (eMethods).^21–23^ None of the selected or created scores utilized AOU, PMBB, or UCLA data for training.

### Calculation of Polygenic Risk Scores

Polygenic scores for each individual in the AOU, PMBB, and UCLA cohorts were calculated using pgsc_calc, which combines individual genotypes with the existing PRS weights.^9,24^ Scores were calculated for each sample using whole-genome sequence data from AOU and imputed genotypes from PMBB and UCLA, and weights from all PGS Catalog scores meeting described criteria, as well as PGS_LDP2Auto and PGS_prscsx. Match rate, or the percent of variants in a weight file that were successfully identified in the genomic data, was recorded.

Heterogeneity of allele frequencies and linkage disequilibrium patterns across populations can influence raw PRS distributions, limiting interpretation and generalizability.^8,25^ To minimize these effects, scores were adjusted using a principal component analysis (PCA)-based method to normalize both mean and variance to the 1000 Genomes + HGDP reference panel (eMethods).^26^ All downstream analysis utilized PCA-normalized scores. These values were accordingly translated into risk percentiles based on a standard normal distribution.

PRS performance in diverse populations depends on representation of genetically similar individuals in the GWASs used to construct the PRS.^15,27^ CAD PRSs vary widely in their use of diverse GWAS data. To characterize the impact of genetic background on PRS performance, population subgroup-stratified sensitivity analyses were conducted. Individuals were assigned to one of six population groups based on their genetic similarity to populations included in the 1000 Genomes + HGDP reference panel: African (AFR), admixed American (AMR), East Asian (EAS), European (EUR), Middle Eastern (MID), and Central/South Asian (CSA). Sensitivity analyses were conducted in the AFR and EUR groups. Population group assignment was implemented using pgsc_calc (eMethods).^28^

### Population-Level Assessment: Identification of equivalently performing polygenic risk scores

The association between each PRS and prevalent CAD was assessed using generalized linear regression models with a logit link, including age and sex as covariates. These covariates were included to minimize ascertainment bias. Genetic principal components were not included in the primary analysis model on the premise that they are already accounted for by PCA-based score normalization. Inclusion of PCs, as well as exclusion of all covariates were both evaluated as sensitivity analyses. The approach to risk model evaluation considered relevant published reporting standards^11,12^: risk score distribution was assessed visually, and PRS effect size was quantified with odds ratios; discrimination and calibration were visualized with calibration plots and ROCs, and quantified using Brier score and area under the receiver operator curve (AUROC). Bayesian methods were used to perform between-model comparisons and identify a set of PRSs with statistically and practically equivalent performances relative to the ‘best’ performing score for each metric (eMethods).

### Individual-Level Polygenic Risk Score Assessment

United States regulatory guidelines state that new clinical tests should be evaluated by assessing the agreement between the results of new test and that of a reference standard.^29^ Scores were therefore evaluated through this lens of agreement to determine how consistently scores with comparable population-level performance assigned individual risk.

First, descriptive statistics were calculated to understand the individual-level distributions of genetic risk estimations provided by equivalently performing scores. Each individual’s average risk percentile was calculated, defined as the mean of percentiles assigned by each score meeting the Region of Practical Equivalence (ROPE) of 0.02, as ‘practical equivalence’ criteria. In addition, we calculated the standard deviation and the ratio of the standard deviation to the mean (i.e., the coefficient of variation (CV)) across scores per individual. Distributions of mean risk percentiles and standard deviations across biobank populations were plotted, and we calculated the population-level median of the individual-level average scores, standard deviations, and CV. Bootstrapped 95% confidence intervals were obtained using the “boot” and “simpleboot” R packages.

Agreement across scores was next assessed using inter-rater reliability. If one considers PRSs with equivalent performance as independent, equally qualified testers of the same phenomenon (in this case, genetic liability for CAD), one can apply tests of inter-rater reliability to determine whether scores that are equivalent are also practically interchangeable. The intraclass correlation coefficient (ICC), which is used as a metric of the consistency of quantitative measurements made by different raters measuring the same quantity, was used to compare PRS percentile as a continuous variable.^30^

Since many tested and proposed clinical applications of PRS use percentile cutoffs to define ‘high-risk’ groups, agreement of binary assignment above/below these thresholds was also assessed by evaluating inter-rater agreement of the categorical classification. This was quantitatively assessed using Light’s Kappa (eMethods).^31^

Pearson’s correlation coefficient (r) of individual risk estimates was calculated for all pairs of scores in the primary cohort.

### Statistical Analysis

All analyses were performed using R (version 4.3.0 in PMBB, Version 4.3.1 in AOU and UCLA). Statistically equivalent scores were defined as those with a less than 95% probability of a positive difference of both Brier score and AUROC. Practically equivalent scores were those with a >95% probability of model performance being ‘practically equivalent’ using a ROPE estimate.^32,33^ An *a priori* practical effect size was set at 0.02, and in sensitivity analyses ranges of 0.01 and 0.005 were considered. For individual-level analysis, Light’s Kappa (κ) and ICC were used to compare categorical and continuous agreement of PRS percentiles. Although both ICC and κ are descriptive measures, guidelines exist to aid interpretation of intermediate values. For both, 0 indicates agreement that is no better than random chance, and 1 indicates perfect agreement. A Kappa between 0.41 and 0.60 can be interpreted as fair agreement; an ICC coefficient between 0.50 and 0.75 is generally interpreted as a fair to good relationship. ^34–37^ Methods are described in further detail in eMaterials.

## Results

### Clinical Characteristics

The AOU study population comprised 17,589 (10.3%) participants with and 153,506 (89.7%) participants without CAD (eResults). 97,265 (63.4%) of those without CAD and 7,682 (43.7%) of those with CAD were female. 100,493 (58.7%) of participants were most genetically similar to a European reference population, 35,590 (20.8%) to an African reference population, 29,805 (17.4%) to an Admixed American reference population, and the remaining to Central/South Asian, East Asian, and Middle Eastern reference populations (ST1, eFigure 2). Those with CAD were older on average (69.3 years versus 54.9) (ST1). Both PMBB and UCLA featured a higher rate of prevalent disease. Of 41,193 PMBB participants, 9,215 (22.4%) had CAD (ST2). Of 53,092 UCLA participants, 12,513 (23.5%) had CAD (ST3). These cohorts are representative of national demographic patterns and are well-powered for PRS assessment and replication, allowing for reliable and generalizable population- and individual-level assessment.

### Population-Level PRS Performance

We first aimed to identify a set of polygenic scores that predicted prevalent CAD in a statistically equivalent way based on population-based performance prediction metrics (**Figure 1**). We obtained total of 50 CAD polygenic scores, consisting of 48 from the PGS catalog and two additional scores constructed specifically for this study (PGS_LDP2Auto and PGS_prscsx, Methods, ST4). The majority of genetic variants included in each score were present for AOU, PMBB, and UCLA participants (ST6-8). We observed that two scores had negative associations with prevalent CAD and were subsequently excluded from downstream analysis (ST9, eFigure3, eResults). When added to a model of prevalent disease that included age and sex as covariates in AOU, the remaining 48 scores were significantly associated with CAD (p<0.05) with odds ratios that ranged from 1.10 (PGS000059) to 1.46 (PGS_LDP2Auto) per standard deviation increase in each PRS (eFigure3, ST9), confirming the expected statistical association of these models.

**Figure 1:**
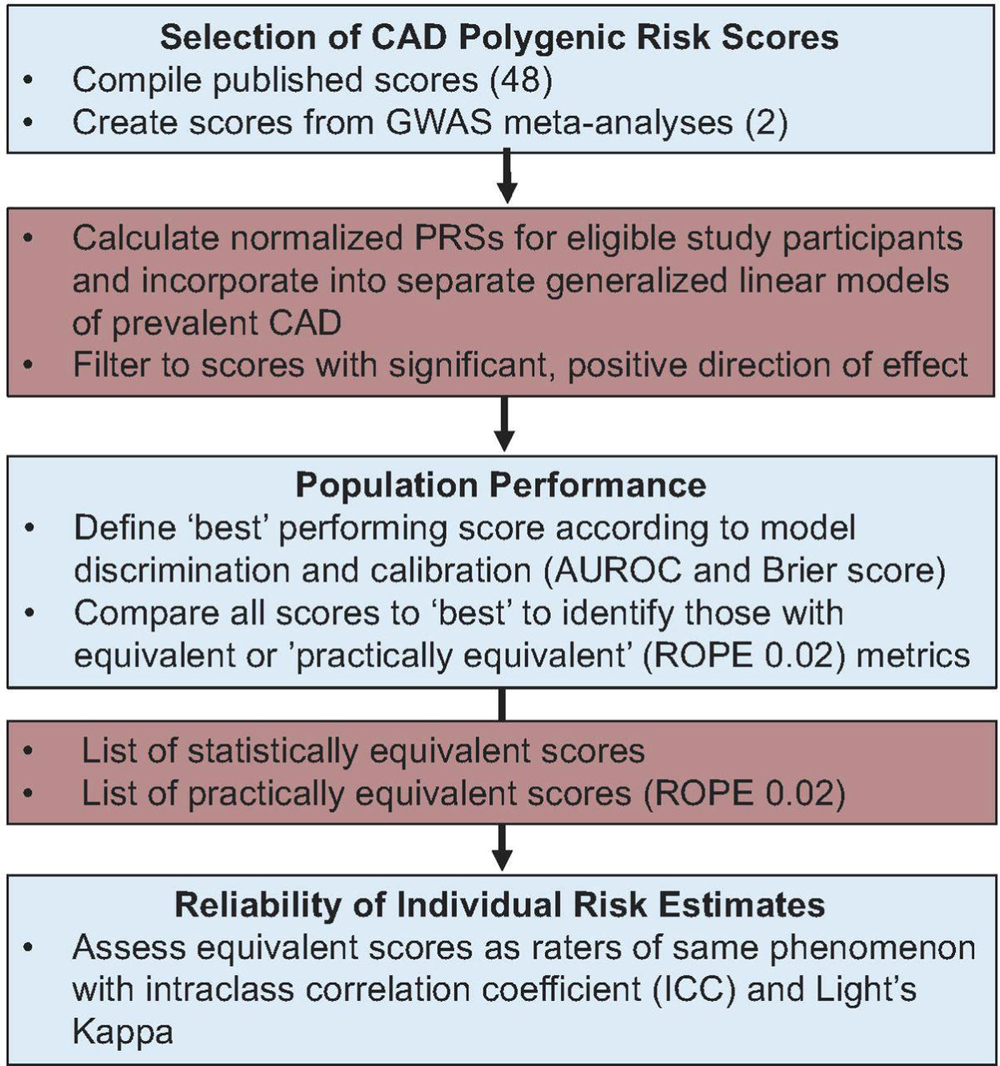
Overview of Approach. 50 polygenic risk scores were calculated for all individuals in the All of Us Research Program, Penn Medicine Biobank, and UCLA ATLAS. In each cohort, the scores with superior population-level performance when included in a model of prevalent disease were identified, and performance metrics were compared with all other scores. Using this to define a list of practically equivalent scores, we considered agreement between risk percentiles assigned individuals by different, ‘equivalent’ scores.

We next evaluated population-based performance measures for a range of prediction metrics. We calculated Brier scores and AUROC for all PRS scores (Methods). We found that the model for the PGS_LDP2Auto score had the best calibration measured by Brier score (0.0825, 95% Credible Interval 0.0829-0.0827), whereas the model including PGS003725 had the best discrimination as measured by AUROC (0.777, 95% Credible Interval 0.777, 0.778) (ST10). To determine the set of score that were practically equivalent, we applied a prespecified primary region of practical equivalence effect size of 0.02 (Methods). All 48 scores had practically equivalent population-level performance (**Figure 2**). Sensitivity analyses that applied more stringent practical effect size margins of 0.01 and 0.005 resulted in 19 and 5 equivalent scores, respectively (ST10). Two scores met our prespecified definition for statistically equivalent performance across both measures of calibration and discrimination. Analyses in PMBB and UCLA revealed similar results, identifying 33 and 48 ROPE 0.02 equivalent scores, respectively (eResults, ST11-14, eFigure 4-7). This result indicates that most CAD PRS are indistinguishable using population-level performance metrics.

**Figure 2:**
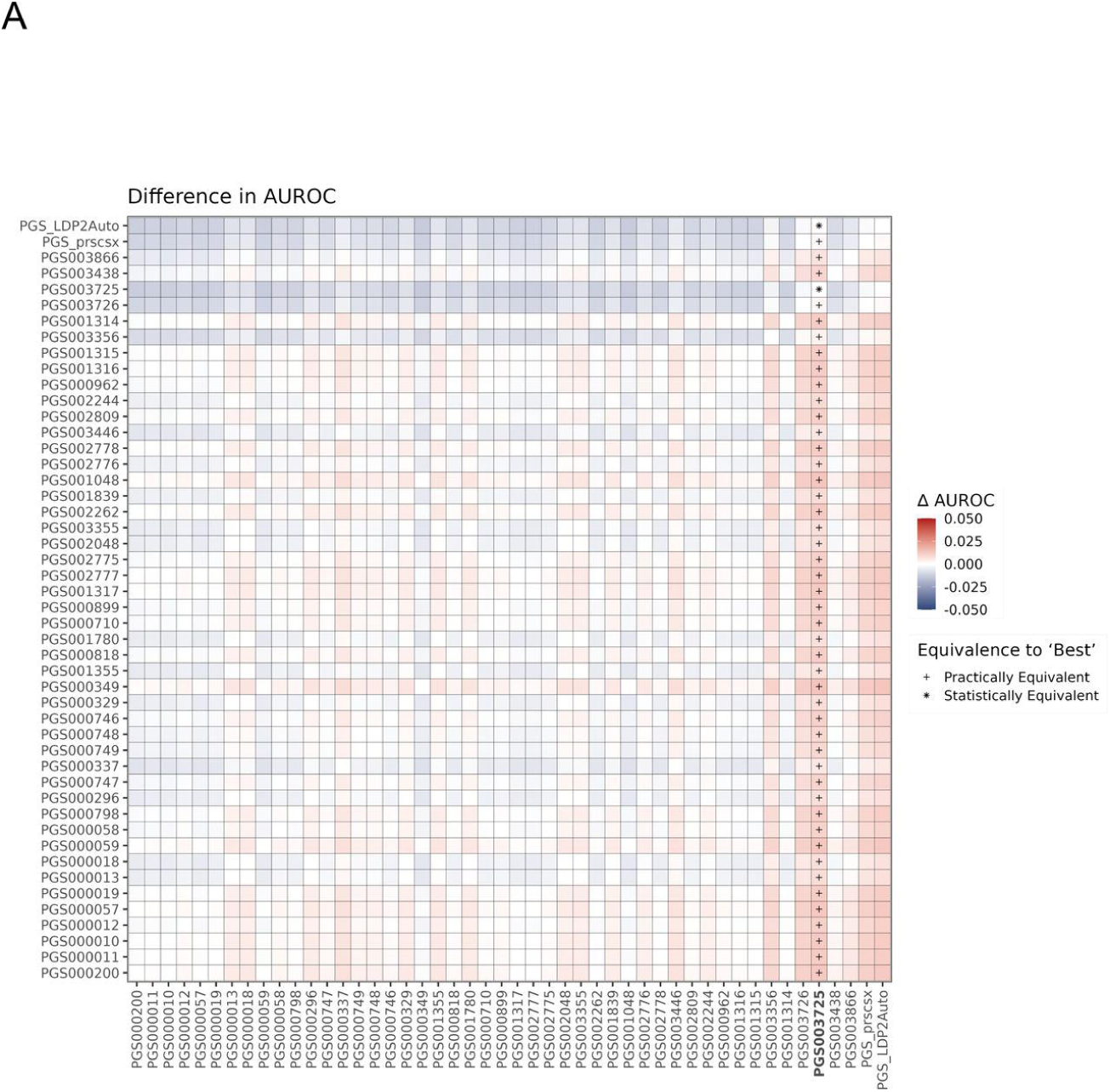
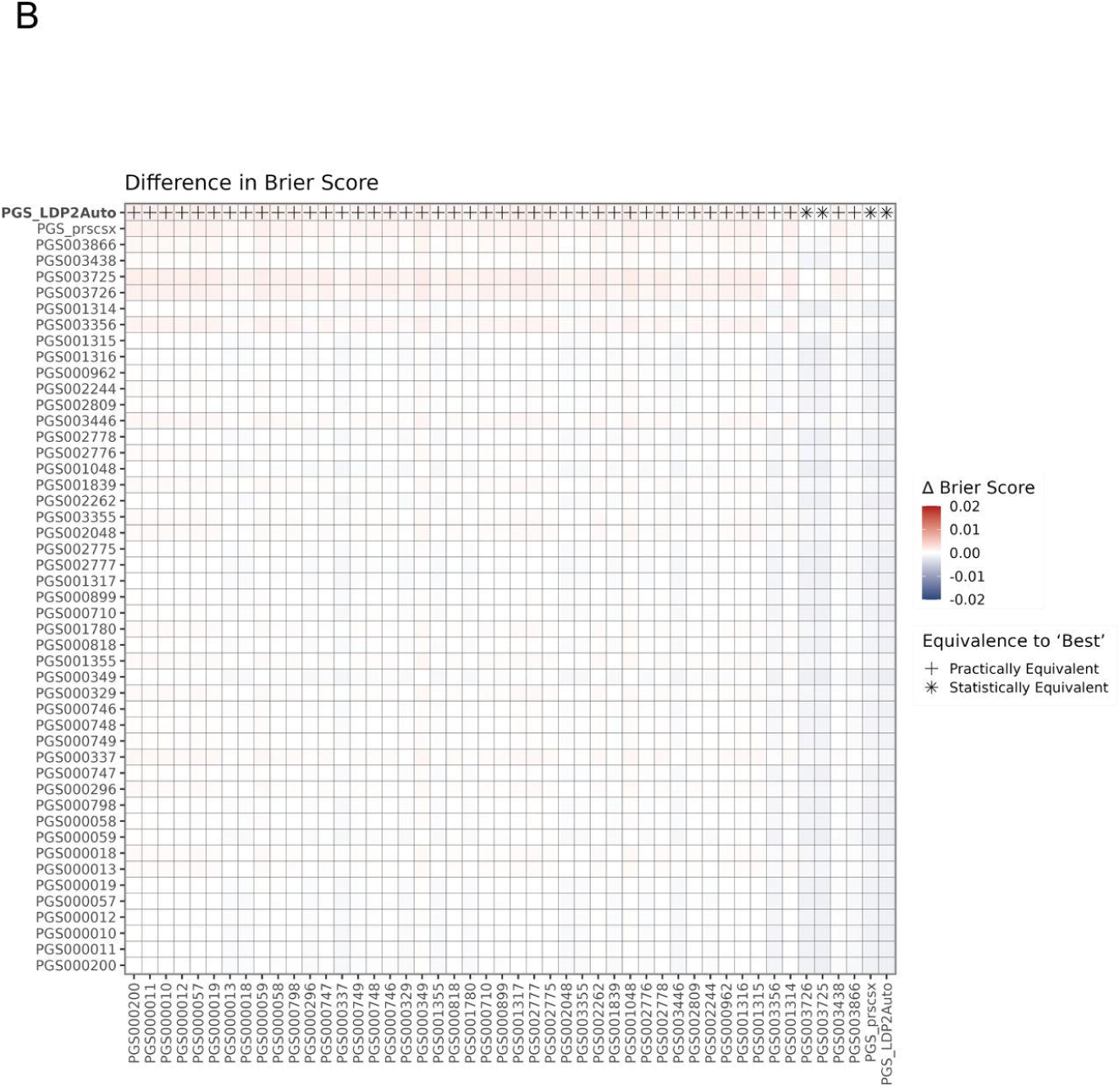
Differences in Polygenic Risk Score AUROCs and Brier Scores in AOU. The mean of the posterior distribution of the difference between the AUROC/Brier Score of each combination of scores is plotted. The score with the ‘best’ model metric is denoted with bold text. Plus signs indicate scores with practically equivalent metrics (ROPE 0.02). Asterisks indicate scores with statistically equivalent metrics. Scores are ordered by year of publication, with PGS_LDPred2Auto and PGS_prscsx (newest) on the right.

Finally, we performed sensitivity analyses in AOU to assess the robustness of the broad population performances of these measures. When stratifying by genetic similarity to continental reference populations or removing model covariates, we observed broadly similarly hierarchies of score performance, but with unique sets of scores meeting equivalence criteria (eResults, ST15-21, eFigure8-15). This result indicates that the best-performing score at the population-level can differ within as well as between cohorts.

### Agreement of Individual PRS Risk Assessments

Having identified a set of CAD PRS that have equivalent population-level metrics, we next evaluated the consistency of individual-level risk estimates. We defined an individual’s average risk percentile as the mean of the PRS percentile risk estimates provided by the 48 scores. Across AOU participants, the median standard deviation of risk percentiles for each individual was 22.94 (22.92, 22.96) and the median coefficient of variation for each individual was 0.504 (0.503, 0.505), providing scale-dependent and -independent evidence of the variability of individual-level estimates (**Figure 3**, eResults). We observed similar in analyses of PMBB and UCLA ATLAS and across sensitivity analyses (eFigure16-21).

**Figure 3:**
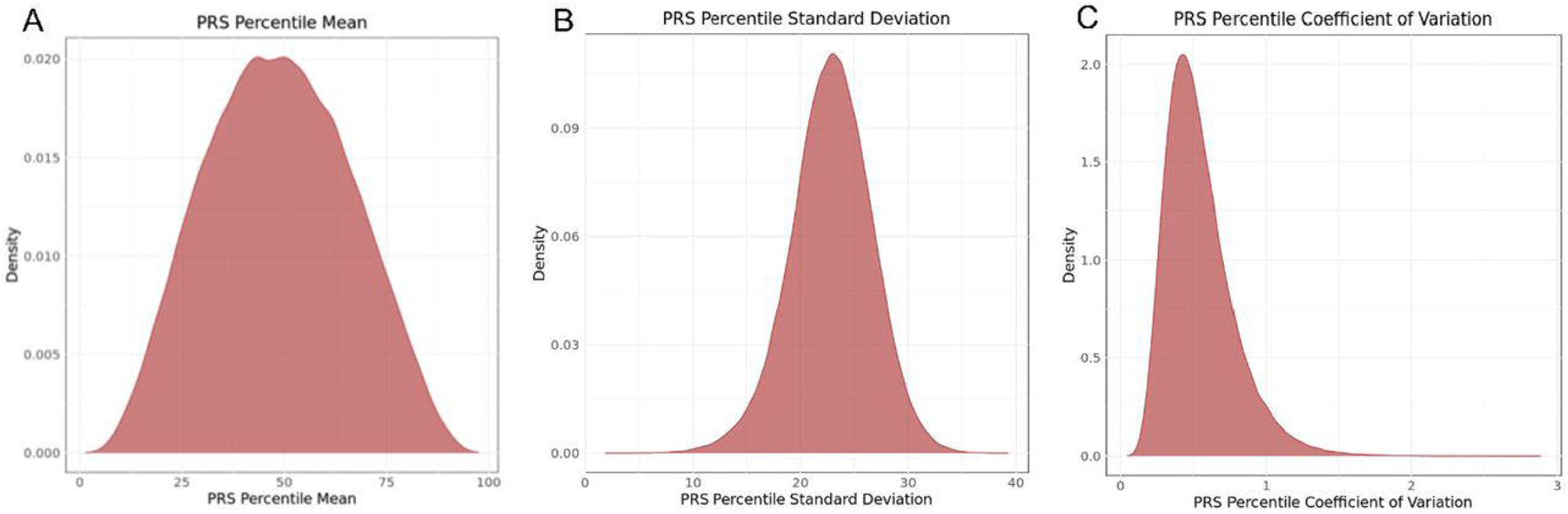
Within-Person Score Concordance in AOU. Concordance of individual score percentiles across all scores meeting practically equivalent ROPE 0.02 criteria in All of Us (48 scores). **A)** Mean individual risk percentile; median 48.38(48.26, 48.49). **B)** Standard Deviation of the mean individual risk percentile; median 22.94 (22.92, 22.96). **C)** Coefficient of Variation; median 0.5039 (0.5028, 0.5052).

Next we examined the distribution of scores in the top / bottom 20^th^ percentiles of risk across all 48 scores. We observed that 84% of individuals had at least one risk estimate above the top 20^th^ risk percentile, and one in the bottom 20^th^ risk percentile (ST22). To help illustrate this individual-level variability, we plotted individual-level risk percentiles for five randomly selected individuals from the 1000 Genomes + HGDP reference population across all practically equivalent scores (**Figure 4**) and designed an interactive web app that allows users to explore CAD PRS percentiles for randomly selected individuals from this population (sabramow.github.io/PRS_Var/). These results empirically highlight how an individual’s estimated risk can differ depending on which score was used.

**Figure 4:**
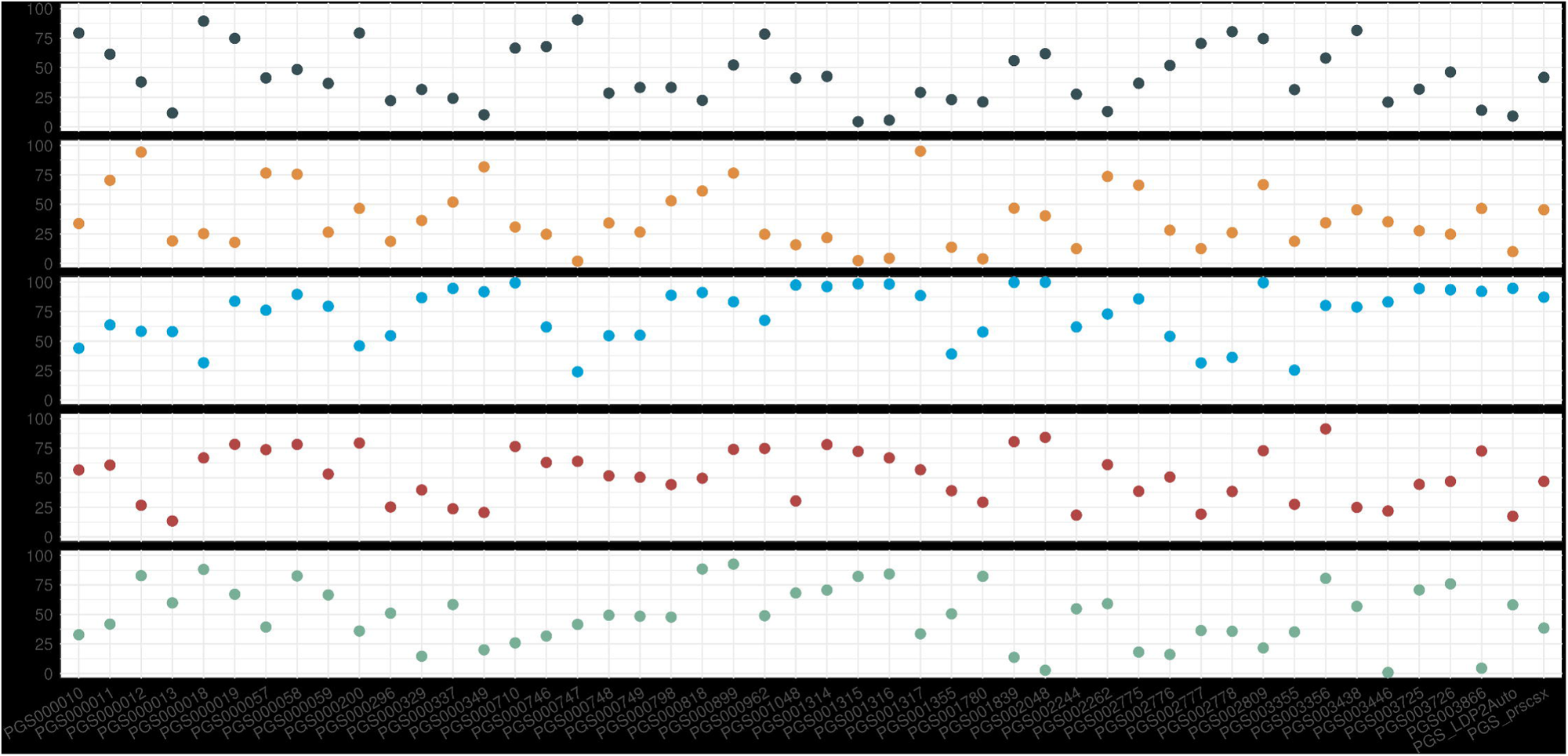
Risk Predictions for Five Randomly Selected Individuals. 48 practically equivalent (defined as ROPE 0.02 in AOU) polygenic risk scores are plotted on the x-axis, in order of year of publication (newest on the right). On the y-axis is polygenic risk score percentile. Color corresponds to a unique randomly selected 1,000 Genomes + HGDP (reference population) participant. Each participant’s risk percentile according to each score is plotted.

We next quantified interrater reliability across scores. Among the 48 practically equivalent scores, ICC was 0.351 (0.349, 0.352) which, according to common interpretation frameworks can be considered poor (**Table 1a**).^35–37^ ICCs for the sets of 5 and 19 scores that met practical equivalence criteria with a ROPE of 0.005 and 0.01 were 0.734 (0.732, 0.736) and 0.555 (0.551, 0.558) respectively. The five ROPE 0.005 equivalent scores included two pairs of scores from the same studies (PGS_prscsx and PGS_LDP2Auto, and PGS003725 and PGS003726). The ICC between the two scores which performed equivalently at the population-level (PGS003725 and PGS_LDP2Auto) of 0.646 (0.643, 0.649) can be considered moderate to good.^35–37^ Because thresholding PRSs to assign individuals into “high-risk” categories is commonly applied, Light’s Kappa was then used to assess whether scores agreed in their risk categorization. Across a range of possible thresholds, performance was almost universally poor but tended to improve with more stringent equivalence thresholds (**Table 1b**, ST22, eResults).

**Table 1a:**
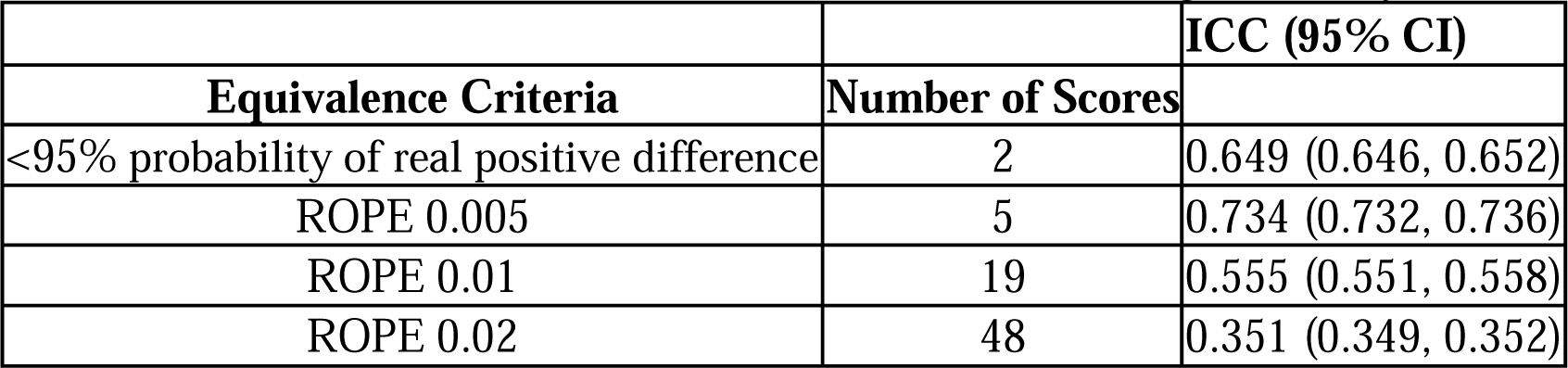
Intraclass Correlation Coefficient. Calculated using a two-way mixed effects model.

|  |  | **ICC (95% CI)** |
| --- | --- | --- |
| **Equivalence Criteria** | **Number of Scores** |  |
| <95% probability of real positive difference | 2 | 0.649 (0.646, 0.652) |
| ROPE 0.005 | 5 | 0.734 (0.732, 0.736) |
| ROPE 0.01 | 19 | 0.555 (0.551, 0.558) |
| ROPE 0.02 | 48 | 0.351 (0.349, 0.352) |

**Table 1b:** Light’s Kappa. Values reflect congruence of scores meeting a denoted equivalence criteria in stratifying individuals above a specified percentile (99, 95, 90, 80, 70, 50).

|  |  | **Light's Kappa** |  |  |  |  |  |
| --- | --- | --- | --- | --- | --- | --- | --- |
| **Equivalence Criteria** | **Number of Scores** | 99 | 95 | 90 | 80 | 70 | 50 |
| <95% probability of real positive difference | 2 | 0.233 | 0.340 | 0.390 | 0.436 | 0.464 | 0.476 |
| ROPE 0.005 | 5 | 0.334 | 0.431 | 0.476 | 0.520 | 0.542 | 0.557 |
| ROPE 0.01 | 19 | 0.182 | 0.267 | 0.310 | 0.357 | 0.381 | 0.401 |
| ROPE 0.02 | 48 | 0.089 | 0.146 | 0.177 | 0.213 | 0.231 | 0.245 |

Next, we repeated our approach in our other settings. In each of these analyses, we used the corresponding sets of equivalent and practically equivalent scores determined for each analysis cohort. We found that metrics and patterns of variability and risk percentile congruence were similar we replicated our analysis pipeline in population-stratified AOU analysis, and in PMBB and UCLA (eResults, ST23-24, ST26-27). These results confirmed that low inter-rater reliability was not exclusive to our primary cohort.

Noting that some pairs of scores seemed to produce more highly correlated individual estimates than others, we considered whether improvements in performance associated with improvements in agreement. For our primary cohort, we produced a heatmap of the correlation coefficients between all score pairs, and also plotted this coefficient by the difference between the performance metrics of the two scores. We observed that correlation between risk estimates provided by pairs of scores varied widely. Two main trends emerged: correlation was highest among pairs of scores derived from the same studies, and also tended to be higher among scores that performed similarly overall and better compared to the rest of the rest of the scores (eFigure 22-23). This trend suggests that using a common data set to generate multiple scores can inflate agreement, and that inter-score agreement may be improving with time and score performance.

## Discussion

In this study, we designed and implemented a framework to evaluate whether CAD PRSs that have similar population level performance provide consistent and reliable assessments of individual-level disease liability. Results from 263,035 individuals across three large-scale biobanks representative of national demographics demonstrate that individual-level estimates of genetic susceptibility to CAD vary considerably across extant PRSs. Although an individual’s genetic risk is fixed from conception, we found that 48 tested scores provided inconsistent individual-level estimates of CAD risk. These scores cannot be distinguished on population-level performance alone, and results of assessments of individual-level agreement call into question the validity of PRSs as clinical tests of the same phenomenon.

A central implication of these results is that PRSs which demonstrate equivalent performance at a population level for CAD may not be considered reliable, interchangeable clinical tests at the individual level. We found that inter-rater reliability estimates of practically equivalent (ROPE 0.02) PRSs were poor, with a high coefficient of variation (50%), and 84% of participants had at least one risk estimate in both the top and bottom quintile. For comparison, The National Cholesterol Education Program recommends that laboratories measure LDL cholesterol with a coefficient of variation of less than 4%.^38,39^ Similar patterns were seen in UCLA and PMBB, and are consistent with a prior report of PRSs applied in the UK Biobank.^40^ Even among smaller sets of scores that were both practically and statistically indistinguishable, inter-rater reliability was not strong.

Tests of inter-rater reliability assume that raters are equally qualified, and therefore the existence of a singular ‘best’ PRS could in theory eschew the need to consider the agreement of different score estimates, as only the ‘best’ score’s should matter. However, multiple factors make this scenario challenging to practically envision and implement. First, objectively identifying a single superior score may not be possible. Implementing our framework for population-level performance assessment, we observed that the ‘best’ score varied by performance metric, cohort, and subpopulation. These conclusions are consistent with prior literature demonstrating that PRS performance varies depending on the environment in which it is assessed, owing to factors such as population structure.^27,41^ While our findings support that larger databases, newer statistical techniques,^42–44^ population-specific focus,^10,45^ and incorporation of susceptibility for ASCVD-related variables^46–48^ do tend to translate to globally better scores, no single score performed best in all analyses, and improvements were marginal (in most cases than our prespecified ROPE of 0.02). A second, related consideration is that score hierarchy is dependent on time. To clinically implement the ‘best’ available score would mean continually updating individual risk predictions, with each new score existing in the context of its predecessors. The degree to which these updates prompt changes in clinical management will reflect the concordance between new and old scores.

Our data indicate a substantial potential for individuals undergoing PRS testing to receive discordant results. There is currently no infrastructure directing patients and providers on how to navigate and reconcile conflicting risk estimates, as the concept of inter-score reliability has been historically absent from ongoing discussions surrounding whether CAD PRSs are ‘ready’ for clinical use.^49,50^ There are also a growing number of proprietary scores which cannot be evaluated through the framework outlined here. Issues of score reliability may be improved with future FDA regulation of laboratory developed tests, ^51^ and our data suggest that population-level score improvement tends to increase – although not yet resolve – inter-rater reliability. Nevertheless, the current availability of multiple PRSs – especially those with indistinguishable or unknown performance – has the potential to undermine and complicate PRS’s clinical validity and reliability, and implementation may lead to confusion and harm.^52^

Taken collectively, our results motivate improvements in the framework with which future CAD PRSs are evaluated, guidelines are issued, and investigation are made that balances a desire for enhancing population-level performance with the pragmatic consideration of the ramifications of the proliferation of scores that disagree in their individual-level risk estimates.

### Limitations

This study should be interpreted in the context of its limitations. First, although we selected scores that were available from the PGS Catalog at the time of analysis, PRSs for CAD are continuously being developed. Second, primary conclusions regarding relative population-level performance of these scores are specific to tested models of prevalent CAD (including population-stratified, and with and without age and sex as covariates). Prevalent disease was chosen as a primary outcome to maximize number of cases and therefore power to detect differences in score performance. Other predictive modeling scenarios have been proposed^53^ and our results do not preclude the possibility that scores can be more conclusively differentiated from these other models. Third, minor differences in sequencing and imputation methodology (which in turn affects match percentage) and phenotype definitions used in UCLA, PMBB, and AOU may account for some of the variability between score performance metrics obtained from each biobank.^54^ Fourth, tests of inter-rater reliability assume that raters (in our case, PRSs) provide independent estimates of risk, but our tested PRSs cannot be considered truly independent raters because in most cases the scores build on a core set of CAD GWAS data. Although a lack of true independence does limit the formal validity of our inter-rater reliability metrics, the effect should be to bias the test statistic towards a higher degree of agreement/correlation, and thus only strengthens our interpretation of substandard inter-rater reliability.

### Conclusions and Relevance

When tested across three diverse biobanks, CAD PRSs that performed similarly at the population level demonstrated highly variable individual-level estimates of risk. Approaches to clinical implementation of CAD PRSs need to consider the potential for incongruent individual risk estimates from otherwise indistinguishable scores.

## Data Availability

Upon publication, the two original polygenic risk scores for CAD will be deposited on the Polygenic Score Catalog. R code for a generalizable score comparison pipeline will be available at https://github.com/Sabramow/PRS_Var. Summary statistics from the genome-wide association study meta-analysis will be available upon reasonable request.

## Acknowledgments

This work would not be possible without the All of Us Research Program, Penn Medicine Biobank, and UCLA ATLAS Precision Health biobank:

We gratefully acknowledge the All of Us Program and its participants. The All of Us Research Program is supported by the National Institutes of Health, Office of the Director: Regional Medical Centers: 1 OT2 OD026549; 1 OT2 OD026554; 1 OT2 OD026557; 1 OT2 OD026556; 1 OT2 OD026550; 1 OT2 OD 026552; 1 OT2 OD026553; 1 OT2 OD026548; 1 OT2 OD026551; 1 OT2 OD026555; IAA #: AOD 16037; Federally Qualified Health Centers: HHSN 263201600085U; Data and Research Center: 5 U2C OD023196; Biobank: 1 U24 OD023121; The Participant Center: U24 OD023176; Participant Technology Systems Center: 1 U24 OD023163; Communications and Engagement: 3 OT2 OD023205; 3 OT2 OD023206; and Community Partners: 1 OT2 OD025277; 3 OT2 OD025315; 1 OT2 OD025337; 1 OT2 OD025276.

The PMBB is supported by Perelman School of Medicine at University of Pennsylvania, a gift from the Smilow family, and the National Center for Advancing Translational Sciences of the National Institutes of Health under CTSA award number UL1TR001878.

We gratefully acknowledge the support of the Institute for Precision Health, participating patients from the UCLA ATLAS Precision Health Biobank, UCLA David Geffen School of Medicine, UCLA Clinical and Translational Science Institute grant number UL1TR001881, and UCLA Health.

We also thank the participants and researchers of the Million Veterans Program, FinnGen, China Kadoorie Biobank, Biobank Japan, MGBB, CardiogramplusC4D, the UK Biobank, and all other biobanks and consortia whose available GWAS data were meta-analyzed for this study. Additionally, much of this work leveraged the robust tools and platform provided by the PGS Catalog, and we thank its creators as well as the researchers who have published their scores to it.

## Disclosures and conflicts of interest

S.A.A. is supported by the Sarnoff Cardiovascular Research Foundation. K.B. is funded by a NIH T32 grant. B.F.V. is grateful for support for the work from the NIH/NIDDK (DK126194). M.G.L. received support from the Institute for Translational Medicine and Therapeutics of the Perelman School of Medicine at the University of Pennsylvania, the NIH/NHLBI National Research Service Award postdoctoral fellowship (T32HL007843), the Measey Foundation, and Doris Duke Foundation (Award 2023-0224). He has received research funding to the institution from Myome to study a CAD PRS, unrelated to this work. S.M.D. was supported by the US Department of Veterans Affairs Clinical Research and Development Award IK2-CX001780. This publication does not represent the views of the Department of Veterans Affairs or the United States Government. S.M.D. received research support from RenalytixAI and Novo Nordisk, outside the scope of the current research, and is named as a coinventor on a Government-owned US Patent application related to the use of genetic risk prediction for venous thromboembolic disease and for the use of PDE3B inhibition for preventing cardiovascular disease, both filed by the US Department of Veterans Affairs in accordance with Federal regulatory requirements.

SAA had full access to PMBB and AOU data used in the study and takes responsibility for the integrity of the data and the accuracy of the data analysis. KB had full access to UCLA ATLAS data used in the study and takes responsibility for the integrity of the data and the accuracy of the data analysis.

The Penn Medicine BioBank is approved by the University of Pennsylvania Institutional Review Board. Patient Recruitment and Sample Collection for Precision Health Activities at UCLA is an approved study by the UCLA Institutional Review Board (UCLA IRB) IRB#17–001013.

## Data Sharing Statement

Upon publication, the two original polygenic risk scores for CAD will be deposited on the Polygenic Score Catalog. R code for a generalizable score comparison pipeline will be available at https://github.com/Sabramow/PRS_Var. Summary statistics from the genome-wide association study meta-analysis will be available upon request.

