## Supplement for "Population Performance and Individual Agreement of Coronary Artery Disease Polygenic Risk Scores"

**ONLINE SUPPLEMENTAL CONTENT**

eMethods	3

Secondary Study Populations	3

Selection of Published CAD PRS Weights	3

CAD Meta-Analysis	3

Creation of Polygenic Risk Score Weights	3

PCA-Based Score Normalization	3

Ancestry Classification	4

Population-Level Assessment	4

Tests of Inter-Rater Reliability	4

eResults	4

Selection of Study Population	4

Evaluation of Polygenic Risk Scores	5

Stratified Population-Level PRS Performance in AOU	5

Population-Level PRS Performance in PMBB	5

Population-Level PRS Performance in UCLA ATLAS	5

Population-Level PRS Performance in AOU, in Model without Covariates	5

Population-Level PRS Performance in AOU, in Model Including Age, Sex, and Principal Components	6

Inter-Rater Reliability of Practically Equivalent Scores	6

eFigures	6

eFigure 1: Flowchart summarizing CAD multi-population meta-analysis.	6

eFigure 2: Principal Component Plots for A) All of Us, B) Penn Medicine Biobank, and C) UCLA ATLAS	7

eFigure 3: Forest Plot of Odds Ratios – AOU	8

eFigure 4: Forest Plot of Odds Ratios – PMBB	9

eFigure 5: Distribution of Polygenic Risk Score Brier Scores and AUROCs in PMBB.	10

eFigure 6: Forest Plot of Odds Ratios in UCLA	11

eFigure 7: Distribution of Polygenic Risk Score Brier Scores and AUROCs in UCLA ATLAS.	12

eFigure 8: Forest Plot of Odds Ratios – AOU AFR	13

eFigure 9: Distribution of Polygenic Risk Score Brier Scores and AUROCs in AOU - AFR	14

eFigure 10: Forest Plot of Odds Ratios – AOU EUR	15

eFigure 11: Distribution of Polygenic Risk Score Brier Scores and AUROCs in AOU - EUR	16

eFigure 12: Forest Plot of Odds Ratios – AOU, PRS Only	17

eFigure 13: Distribution of Polygenic Risk Score Brier Scores and AUROCs in AOU – PRS Only	18

eFigure 14: Forest Plot of Odds Ratios – AOU with PC Covariates	19

eFigure 15: Distribution of Polygenic Risk Score Brier Scores and AUROCs in AOU with PC Coveriates	20

eFigure 16: Within-Person Score Concordance in PMBB.	21

eFigure 17: Within-Person Score Concordance in UCLA	21

eFigure 18: Within-Person Score Concordance in AOU – AFR	22

eFigure 19: Within-Person Score Concordance in AOU – EUR	22

eFigure 20: Within-Person Score Concordance in AOU – PRS Only	23

eFigure 21: Within-Person Score Concordance in AOU – Add PC Covariates	23

eFigure 22: Pearson r AOU	24

eFigure 23: Score Pair Correlation by Magnitude of Performance Difference - AOU	25

eReferences	27

### **eMethods**

#### Secondary Study Populations

The Penn Medicine Biobank recruits from the Penn Medicine health system and links individuals’ biospecimen data to their electronic healthcare record.^1^ The same CAD phenotype definition and genetic sex criteria used in AOU were applied to 43,623 PMBB volunteers with genotyping data. Age was defined as age upon study enrollment.

The UCLA ATLAS Precision Health Biobank recruits from the UCLA Health system and links individuals genotype data to their EHR.^2,3^ CAD phenotype definitions were pulled from the Chronic condition warehouse definitions for myocardial infarction and ischemic heart disease. Sex was pulled from the demographic information available from EHR data. Age was defined as age at the beginning of EHR data availability (1/1/2013) or age at UCLA for those establishing after 2013.

#### Selection of Published CAD PRS Weights

Score weights associated with experimental factor ontology trait (EFO) codes EFO_0001645 (CAD), EFO_0000612 (myocardial infarction), EFO_1001375 (myocardial ischemia), and EFO_0008586 (non-ST-elevation myocardial infarction) that had been released to the PGS Catalog prior to December 15, 2023 were selected.

#### CAD Meta-Analysis

METAL^4^ was used to conduct fixed-effect inverse variance weighted meta-analyses of autosomal GWAS summary statistics from Biobank Japan, China Kadoorie Biobank, FinnGen (Release 9), summary statistics from the Department of Veterans Affairs Million Veteran Program’s Hispanic, Black, and White cohorts, as well as a recent meta-analysis of eight predominantly European cohorts including the UK Biobank and CARDIoGRAMplusC4D Consortium (eFigure1, ST5).^5–9^ There is no known overlap between any of the cohorts composing this meta-analysis.

#### Creation of Polygenic Risk Score Weights

Weights for the “PGS_LDP2Auto” score were created using the “auto” setting of LDpred2 from the ‘bigsnpr’ R package. Meta-analyzed multi-ancestry summary statistic were used, along with a LD reference matrix for 1,444,196 HapMap3+ variants based on European individuals. Weights for “PGS_prscsx” were created with the “auto” setting of PRS-CSx using AFR, EAS, EUR, and AMR summary statistics and corresponding population-specific LD reference panels constructed using 1000 Genomes Project Phase 3 Samples. Both methods use Bayesian approaches to estimate parameters without requiring the use of independent training data. Effective sample size was calculated as 4/((1/n_cases_) + (1/n_controls_)). For both methods, a set seed of 2023 was supplied to ensure reproducibility.

#### PCA-Based Score Normalization

Pgsc_calc’s Z_norm2 approach to PCA-based score normalization was utilized for AOU and PMBB. In this approach, a PCA space is created using reference samples. PRS is modeled as a linear function of the principal components. Residuals are calculated as the difference between the observed and predicted PRS, and then divided by the standard deviation of the residuals. This ensures that the adjusted PRS values have a mean of 0 when considering the influence of genetic ancestry. To address for the variations in spread of PRS, the variance of the residuals is modeled as a function of the PCs, and then is used to normalize the residual PRS. As a result, the variance of the PRS distribution is approximately 1 across populations.

PCA-normalized scoring was also used for UCLA ATLAS. The mean and standard deviation for the first 20 PCs and PGS were calculated and used to derive residuals for each score column. These linear regression derived residuals are used to generate normalized mean and standard deviation of each score.

#### Ancestry Classification

For AOU and PMBB, population group assignment was determined using pgsc_calc’s genetic similarity analysis within its ANCESTRY_ANALYSIS module and tool of its ‘pgscatalog_utils’ package. To identify the population that an individual is most genetically similar too, a RandomForest classifier is trained to classify samples based on their PC-loadings to the most genetically similar population in the provided 1KG + HGDP reference panel.

For UCLA Atlas, population group assignment was inferred as described in previous sources.^13^ Briefly, after quality control steps PCA analysis was performed using FlashPCA 2.0 software with the default settings on all individuals from UCLA and the 1000 Genomes phase 3 dataset. K-nearest neighbors clustering algorithm was applied to group individuals into European, African, American, East Asian and South Asian genetic ancestry and assign individuals to the group with >0.50 cluster membership. Individuals not matched to a cluster with this method were considered unclassified.

#### Population-Level Assessment

Population-level assessment began by restricting to scores that, in a model, had a positive, statistically significant (p<0.05) association with CAD. Resampling using V-fold cross-validation with 6 repeats of 10 folds was used to compute Brier scores and AUROCs for each model. Posterior distributions of these parameters using the ‘perf_mod’ function of the ‘tidyposterior’ R package, which fits resampling statistics using a generalized linear model with a Gaussian error and identity link as well as random effect terms, were generated.^14^ ‘Perf_mod’ was run with 4 chains for 8000 iterations including 1000 discarded warmup iterations. The posterior distributions of the difference between parameters was calculated using the ‘contrast_models’ function, and the model with the ‘best’ performance in each metric was identified.

The posterior distributions of the difference between the metrics of the ‘best’ model and that of every other model were subsequently assessed. For both AUROC and Brier score, the smaller value was subtracted from the larger value.

Two criteria were established for comparing model performance. First, the most stringent equivalence criteria defined statistically equivalent scores as those for which there was a less than 95% probability of a real positive difference of both Brier score and AUROC. The second tested for a >95% probability of model performance being ‘practically equivalent’ using a Region of Practical Equivalence (ROPE) estimate.^15,16^ An *a priori* practical effect size was set at 0.02, and in sensitivity analyses ranges of 0.01and 0.005 were considered.

#### Tests of Inter-Rater Reliability

Inter-rater reliability analyses were performed using the “irr” R package.^17^ ICC was calculated via two-way tests of agreement. Light’s Kappa was used to quantitatively assess the agreement of scores in assigning an individual to above the 50^th^, 70^th^, 80^th^, 90^th^, 95^th^, and 99^th^ percentile. This test computes the arithmetic mean Cohen’s kappa calculated for all ‘rater’ pairs.^18^ A kappa statistic of 1 indicates perfect agreement, and 0 indicates agreement at the level expected of random chance.

### **eResults**

#### Selection of Study Population

PRSs were calculated for all 237,568 All of Us participants with available whole genome sequencing data. Of those, 175,099 had available information allowing for determination of age and CAD status. Excluding the 4,004 individuals without a clear XX or XY genetic sex resulted in a final study population of 171,095 individuals.

PRSs were calculated for all 43,573 Penn Medicine Biobank participants with available imputed genotyping data. Filtering as described above resulted in a study population of 41,193 individuals.

PRSs were calculated for all 54,202 individuals with available imputed genotyping data in UCLA ATLAS Precision Health Biobank. Of these, 28 were dropped due to unavailable EHR sex. 1082 were dropped due to insufficient phenotype information (<1 year follow time) leaving 53,092 for the final analysis.

#### Evaluation of Polygenic Risk Scores

PGS000116 was excluded from downstream analysis because it had a negative association with CAD in AOU, PMBB, and UCLA. This has been previously reported.^19^

PGS003727 had a negative association with CAD in AOU and PMBB. PGS003727 had a positive association with CAD likely due to version inconsistency; the PGS Catalog score file version used to generate results in AOU and PMBB (published 07/14/2023) was replaced on 12/15/2023, and the second version was used in UCLA. The score file was updated for a third time on 05/22/2024.

#### Stratified Population-Level PRS Performance in AOU

Restricting the analysis pipeline to AOU participants most genetically similar to an African reference population, 41 scores had a significant (p<0.05) positive association with prevalent CAD when included in a model that included age and sex as covariates. PRS odds ratios ranged from 1.0 (PGS00011) to 1.17 (PGS_prscsx) (ST10, eFigure 8). The model including PGS_prscsx had the best calibration as determined by the lowest Brier Score (0.085) and highest AUROC (0.72). The ‘best’ (lowest) Brier score in this analysis was higher than the ‘worst’ Brier score in the unstratified analysis, and the ‘best’ (highest) AUROC in this analysis was lower than the ‘worst’ AUROC in the unstratified analysis. All scores were practically equivalent for population-level performance using a practical effect size of 0.02; this was still true when a more stringent ROPE of 0.01 was considered (eFigure 9, ST17). Four scores had statistically equivalent model performance as PGS_prscsx.

Restricting the analysis pipeline to AOU participants most genetically similar to a European reference population, 48 scores had a significant (p<0.05) positive association with prevalent CAD when included in a model that included age and sex as covariates. PRS odds ratios ranged from 1.08 (PGS000349) to 1.67 (PGS_LDP2Auto) (ST10, eFigure 10). The model including PGS_LDP2Auto had the best calibration as determined by the lowest Brier Score (0.09) and highest AUROC (0.79)(eFigure 11). 37 scores were practically equivalent for population-level performance using a practical effect size of 0.02 (ST16). PGS003726 had statistically equivalent model performance as PGS_LDP2Auto.

#### Population-Level PRS Performance in PMBB

In PMBB, PRS odds ratios ranged from 1.056 (PGS000349) to 1.596 (PGS_003725) (eFigure 4, ST13). The model including the PGS003725 score had the best calibration as determined by the lowest Brier score (0.14) and best discrimination as determined by the highest AUROC (0.80). 33 scores were practically equivalent for population-level performance using a practical effect size of 0.02 (eFigure 5). The set of practically equivalent scores decreased in size to 6 and 3 scores when considering practical effect size margins of 0.01 and 0.005, respectively (ST20).

#### Population-Level PRS Performance in UCLA ATLAS

In UCLA ATLAS, PRS OR ranged from 1.034 (PGS000116) to 1.40 (PGS003725) (ST14). The model including PGS003725 had the best calibration as determined by the lowest Brier score (0.142) and best discrimination as determined by the highest AUROC (ST21). 45 scores were practically equivalent for population level performance using a practical effect size of 0.02 (eFigure7). The set of practically equivalent scores decreased in size to 9 and 5 scores when considering practical effect size margins of 0.01 and 0.005 respectively (ST21).

#### Population-Level PRS Performance in AOU, in Model without Covariates

48 scores had a significant (p<0.05) positive association with prevalent CAD when included PRS, but no covariates. PRS odds ratios ranged from 1.04 (PGS001048) to 1.31 (PGS_prscsx and PGS_LDP2Auto) (ST12, eFigure 12). The model including PGS_prscsx had the best calibration as determined by the lowest Brier Score (0.092) and PGS_LDP2Auto had the best discrimination as determined by the highest AUROC (0.57). These two scores were statistically equivalent. Six scores were practically equivalent for population-level performance using a practical effect size of 0.02 (ST19, eFigure 13).

#### Population-Level PRS Performance in AOU, in Model Including Age, Sex, and Principal Components

Including the first five genetic principal components in the model did not change the results of the analysis of population-level performance. Raw score performance metrics were virtually identical, and the list of scores meeting each set of equivalence criteria were the same (ST11,18, eFigure 14-15).

#### Inter-Rater Reliability of Practically Equivalent Scores

Inter-rater reliability was also tested when a more stringent ROPE threshold of 0.005 or 0.01 was used to define practically equivalent scores. Although reliability metrics generally worsened as equivalence stringency – and number of scores deemed equivalent – decreased, there were a few exceptions. Notably, this phenomenon was observed when the smaller set of scores were derived from distinct data sets and the larger set of scores included at least one pair of scores derived from the same genetic data. For example, agreement between the five scores meeting ROPE 0.005 practical equivalence criteria was slightly higher than between the two scores that were statistically equivalent (ST22-23). The five scores included two pairs of scores derived from the same study and summary statistics (PGS_LDPred2Auto and PGS_prscsx, and PGS003725 PGS003726).

### **eFigures**

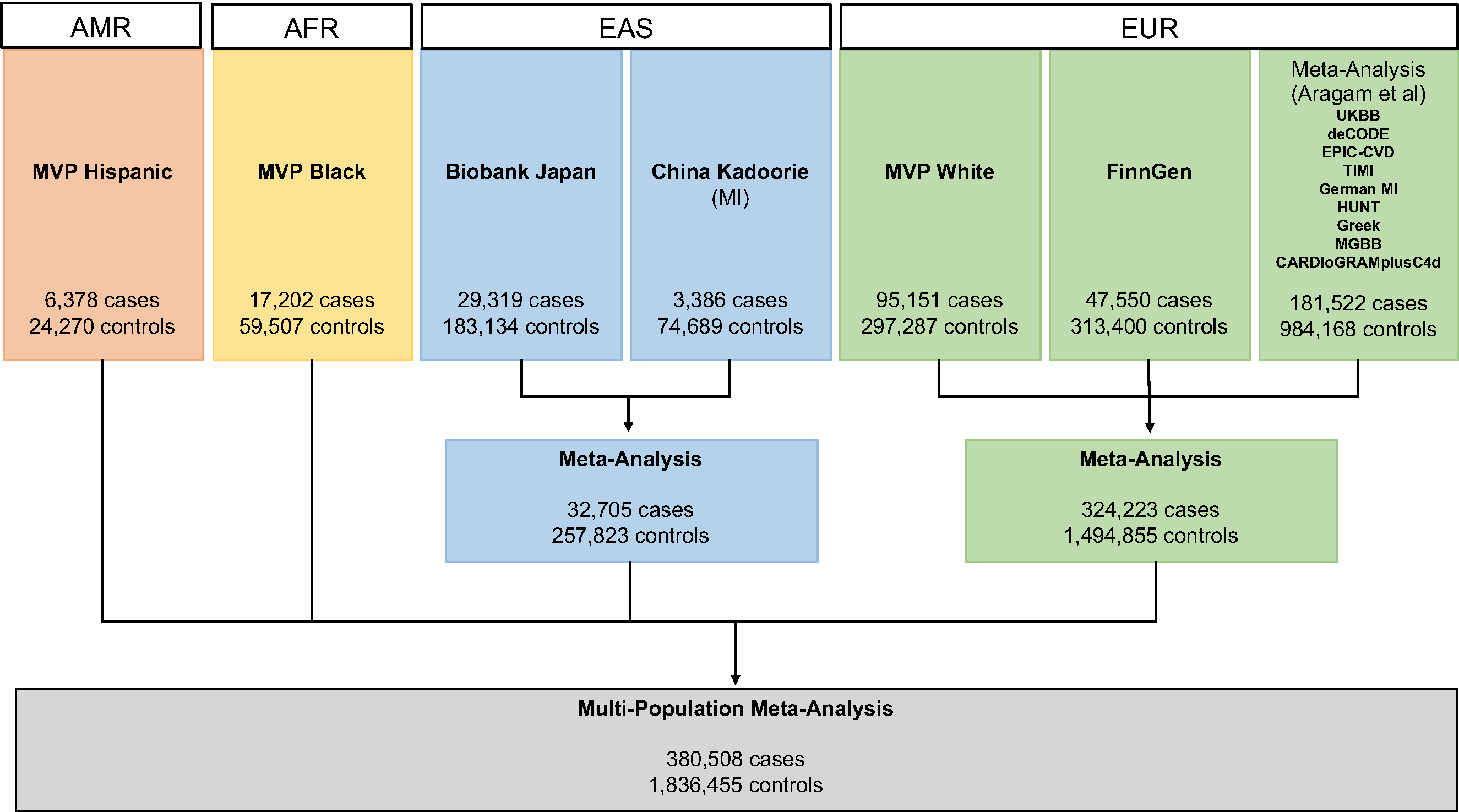

#### **eFigure 1**: **Flowchart summarizing CAD multi-population meta-analysis.**

Study details are in ST1.

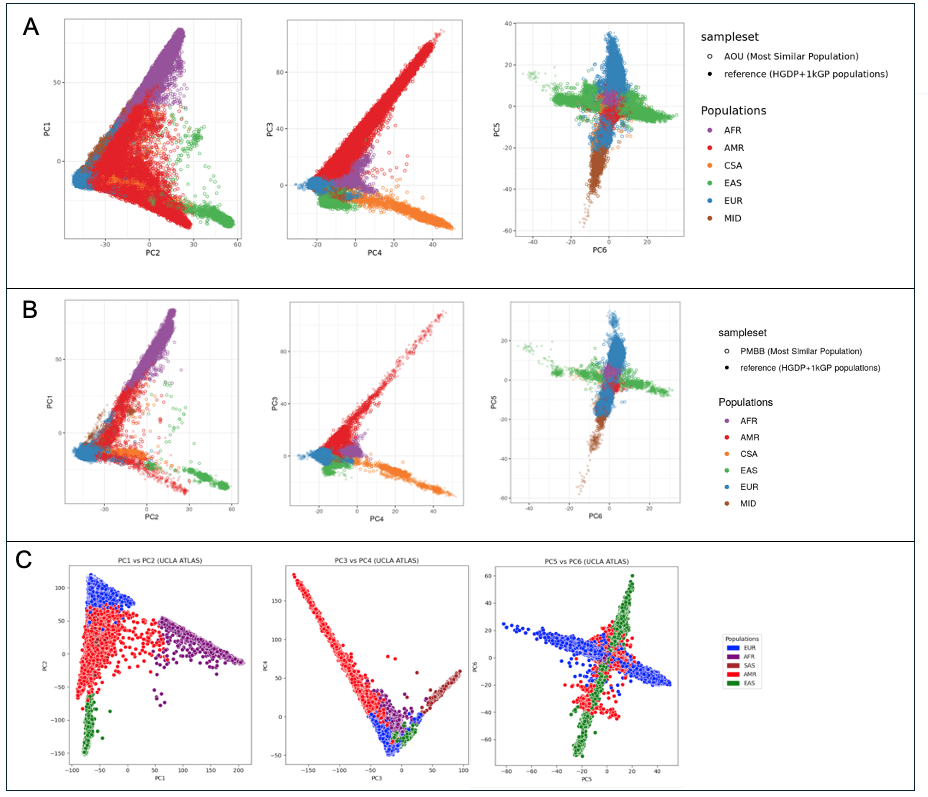

#### **eFigure 2: Principal Component Plots for A) All of Us**, **B) Penn Medicine Biobank,** and **C) UCLA ATLAS**

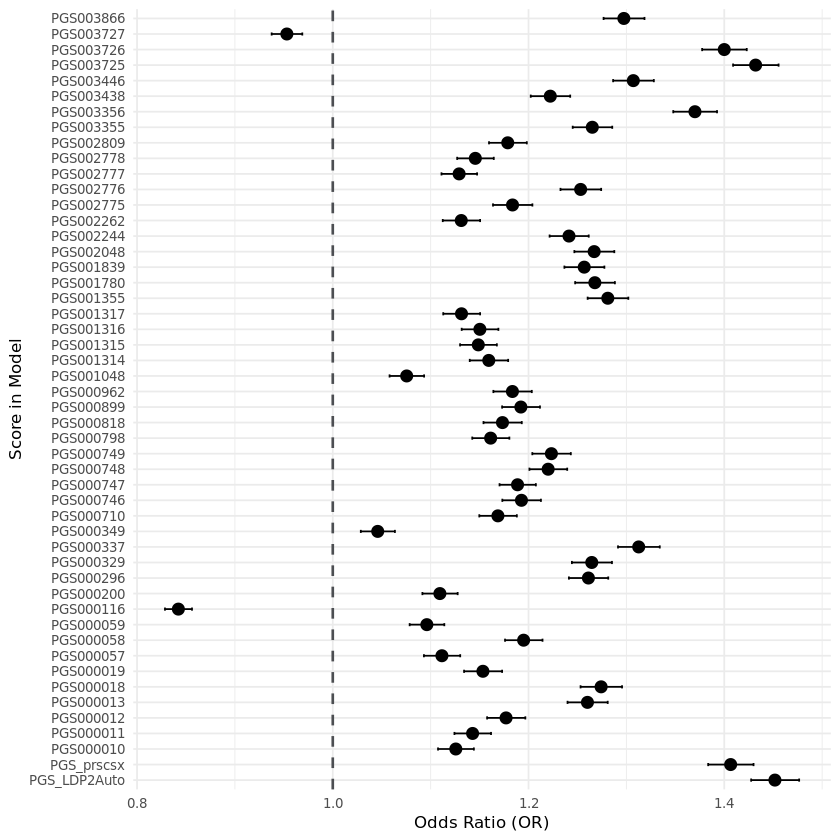

#### **eFigure 3: Forest Plot of Odds Ratios – AOU**

**Forest Plot of Odds Ratios for 50 PRSs in a Model of Prevalent CAD, Including Age and Sex as Covariates in All of Us**

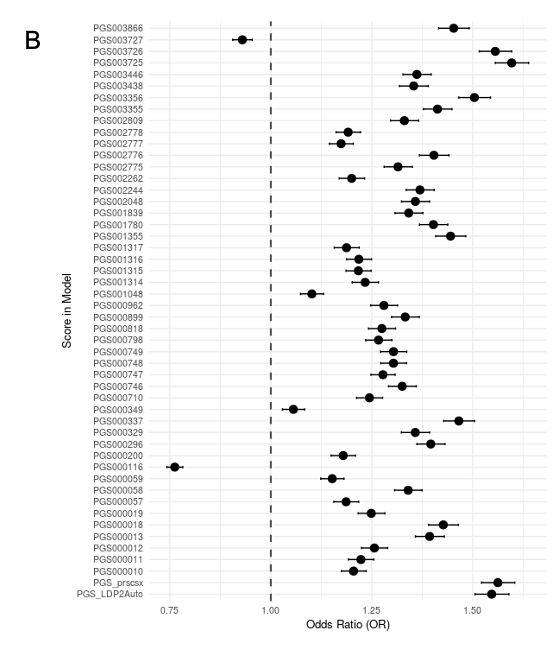

#### **eFigure 4: Forest Plot of Odds Ratios – PMBB**

**Forest Plot of Odds Ratios for 50 PRSs in a Model of Prevalent CAD, Including Age and Sex as Covariates** **in Penn Medicine Biobank**

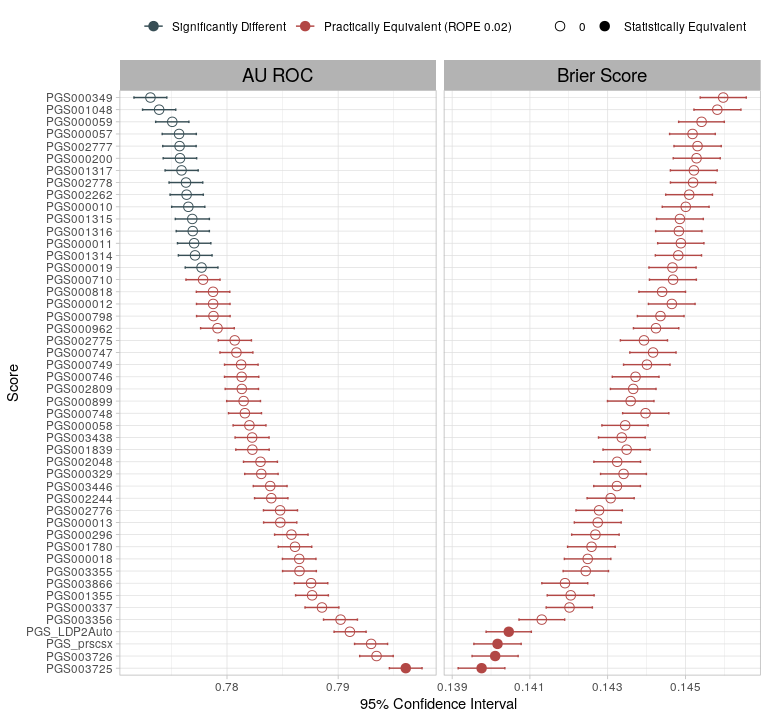

#### **eFigure 5: Distribution of Polygenic Risk Score Brier Scores and AUROCs in PMBB.**

Estimates and 95% confidence intervals are plotted. Scores in red are practically equivalent using a ROPE of 0.02. Scores with filled in circles have <95% probability of a real positive difference (statistically equivalent). Values are in ST20.

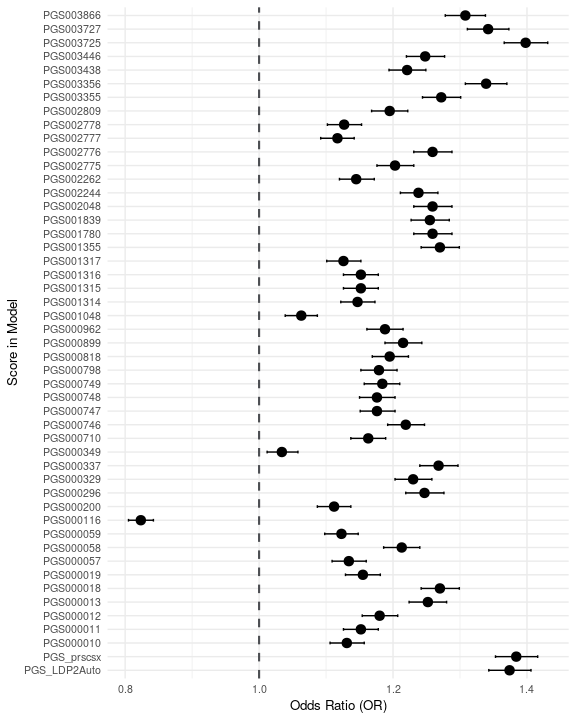

#### **eFigure 6: Forest Plot of Odds Ratios in UCLA**

**Forest Plot of Odds Ratios for 49 PRSs in a Model of Prevalent CAD, Including Age and Sex as Covariates** **in Penn Medicine Biobank**

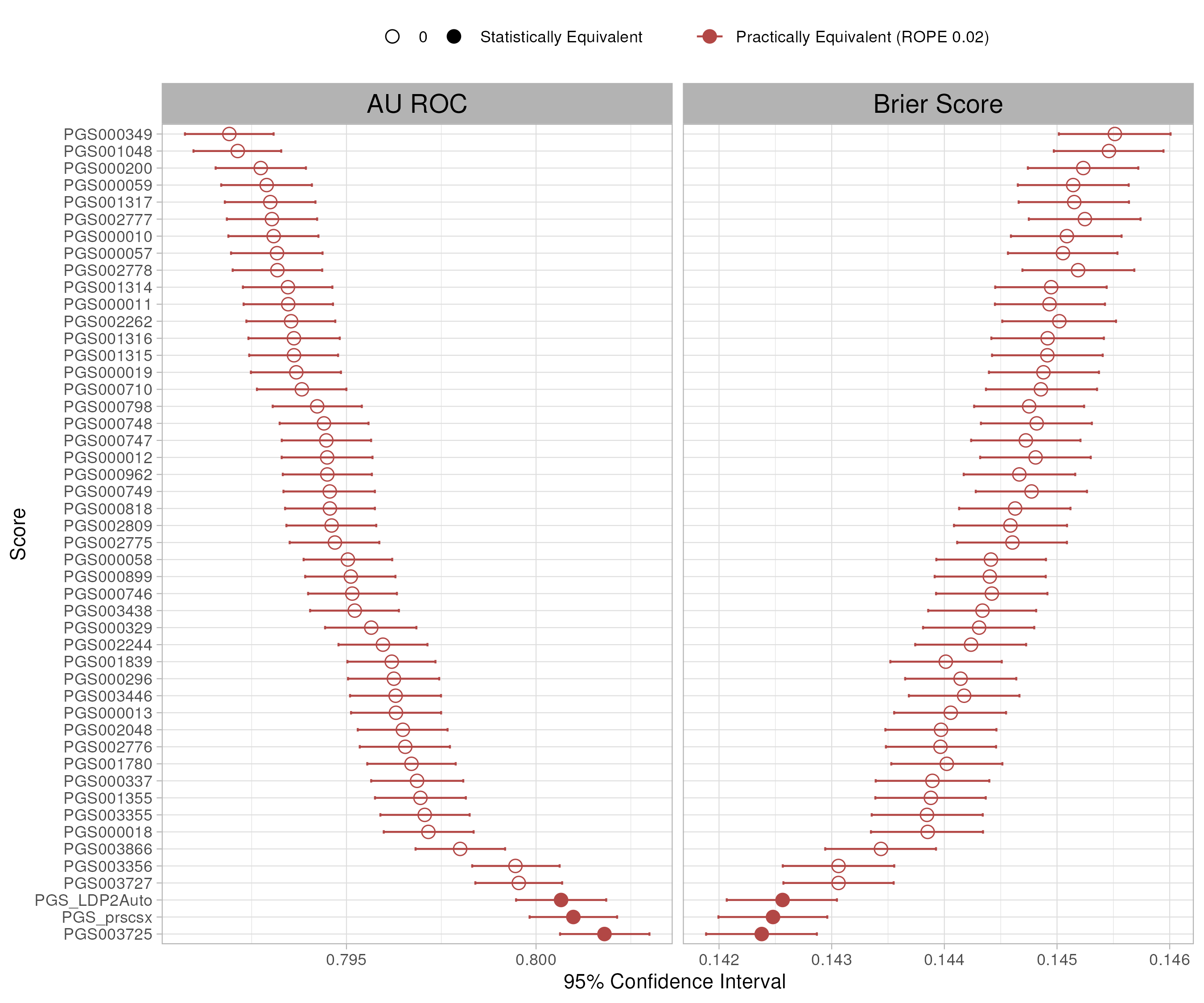

#### **eFigure 7: Distribution of Polygenic Risk Score Brier Scores and AUROCs in UCLA ATLAS.**

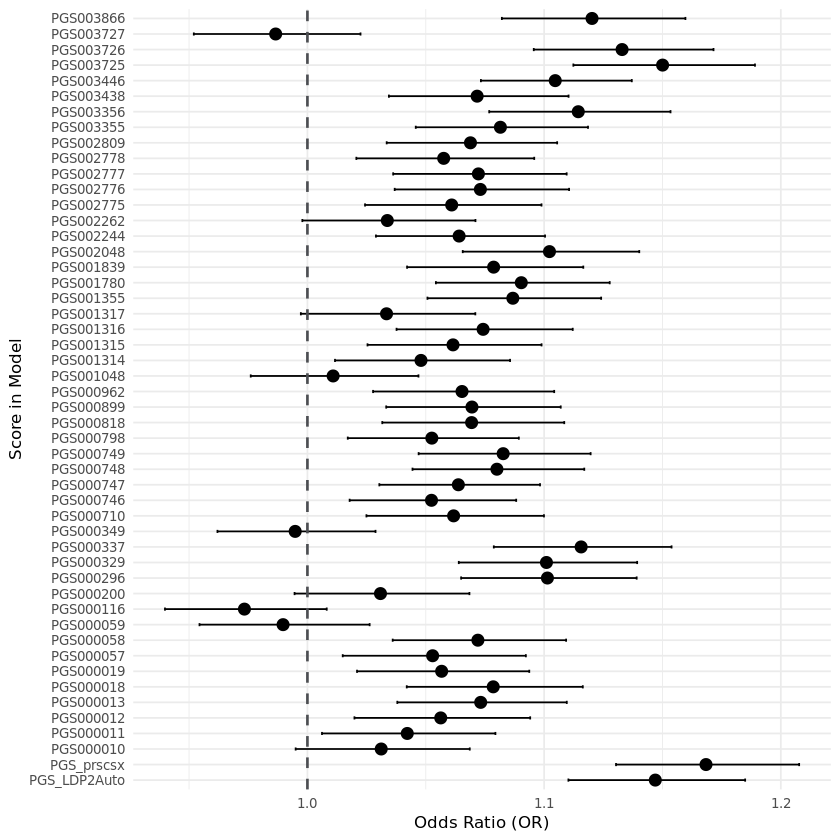

#### **eFigure 8: Forest Plot of Odds Ratios – AOU AFR**

**Forest Plot of Odds Ratios for 50 PRSs in a Model of Prevalent CAD, Including Age and Sex as Covariates in All of Us Individuals most Genetically Similar to an African Reference Population.** Estimates and 95% confidence intervals are plotted. Scores in red are practically equivalent using a ROPE of 0.02. Scores with filled in circles have <95% probability of a real positive difference (statistically equivalent). Values are in ST10.

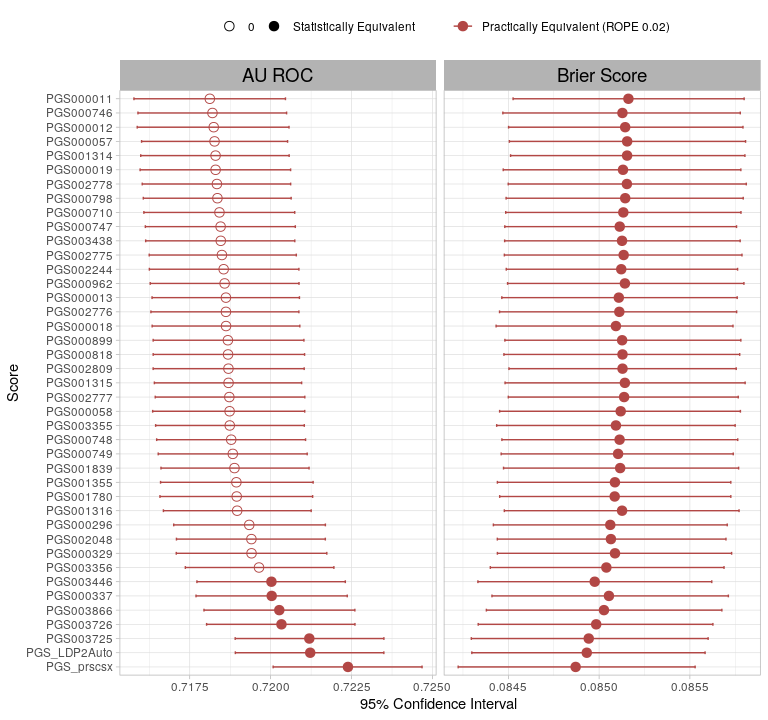

#### **eFigure 9: Distribution of Polygenic Risk Score Brier Scores and AUROCs in AOU - AFR**

**Distribution of Polygenic Risk Score Brier Scores and AUROCs in AOU including Age and Sex as Covariates, in All of Us Individuals most Genetically Similar to an AFR Reference Population.** Estimates and 95% confidence intervals are plotted. Scores in red are practically equivalent using a ROPE of 0.02. Scores with filled in circles have <95% probability of a real positive difference (statistically equivalent). Values are in ST17.

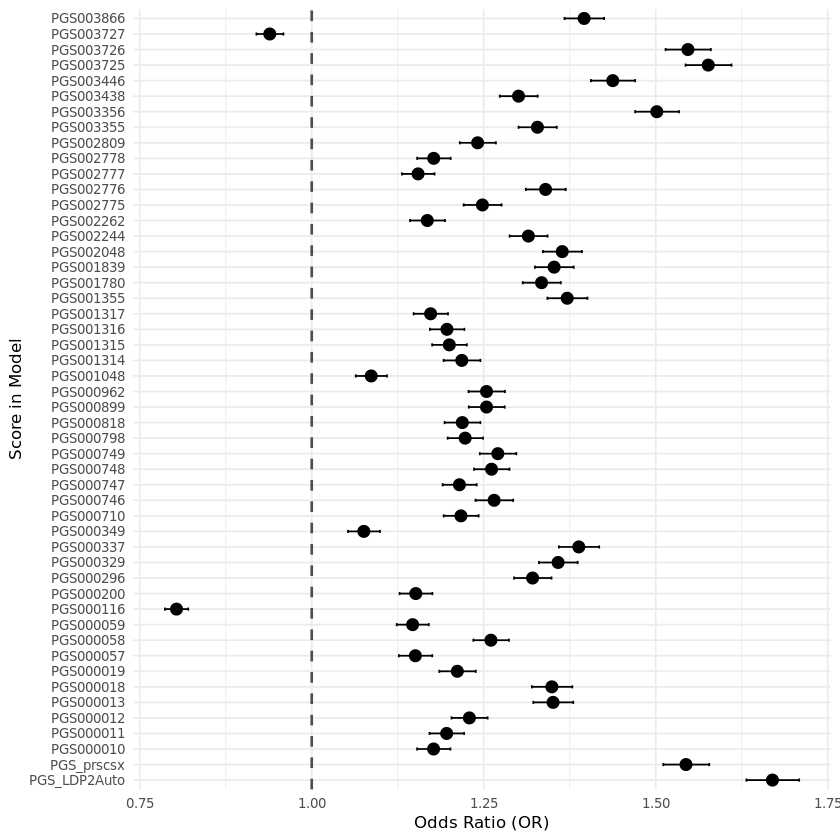

#### **eFigure 10: Forest Plot of Odds Ratios – AOU EUR**

**Forest Plot of Odds Ratios for 50 PRSs in a Model of Prevalent CAD, Including Age and Sex as Covariates in All of Us Individuals most Genetically Similar to a European Reference Population**

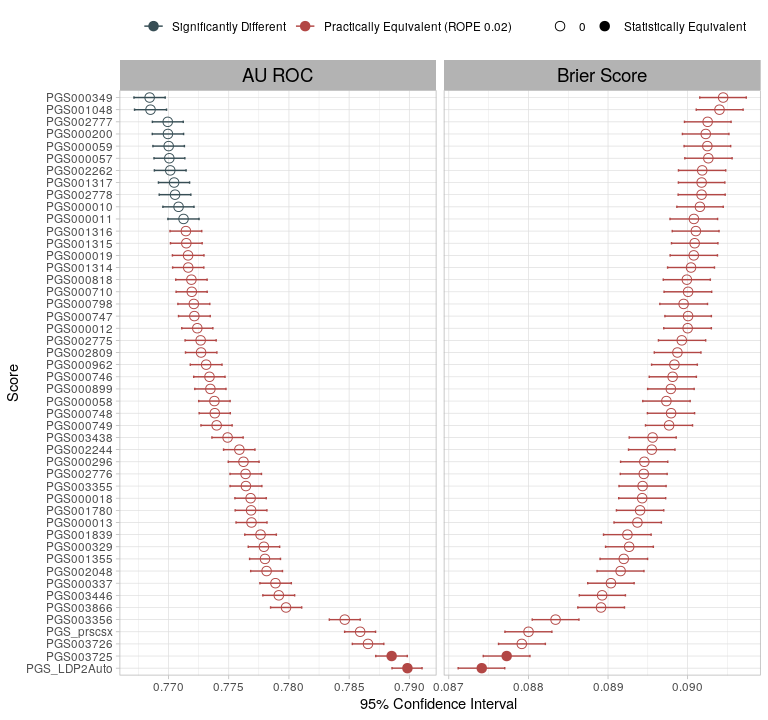

#### **eFigure 11: Distribution of Polygenic Risk Score Brier Scores and AUROCs in AOU - EUR**

**Distribution of Polygenic Risk Score Brier Scores and AUROCs in AOU, including Age and Sex as Covariates, in All of Us Individuals most Genetically Similar to a European Reference Population.** Estimates and 95% confidence intervals are plotted. Scores in red are practically equivalent using a ROPE of 0.02. Scores with filled in circles have <95% probability of a real positive difference (statistically equivalent). Values are in ST16.

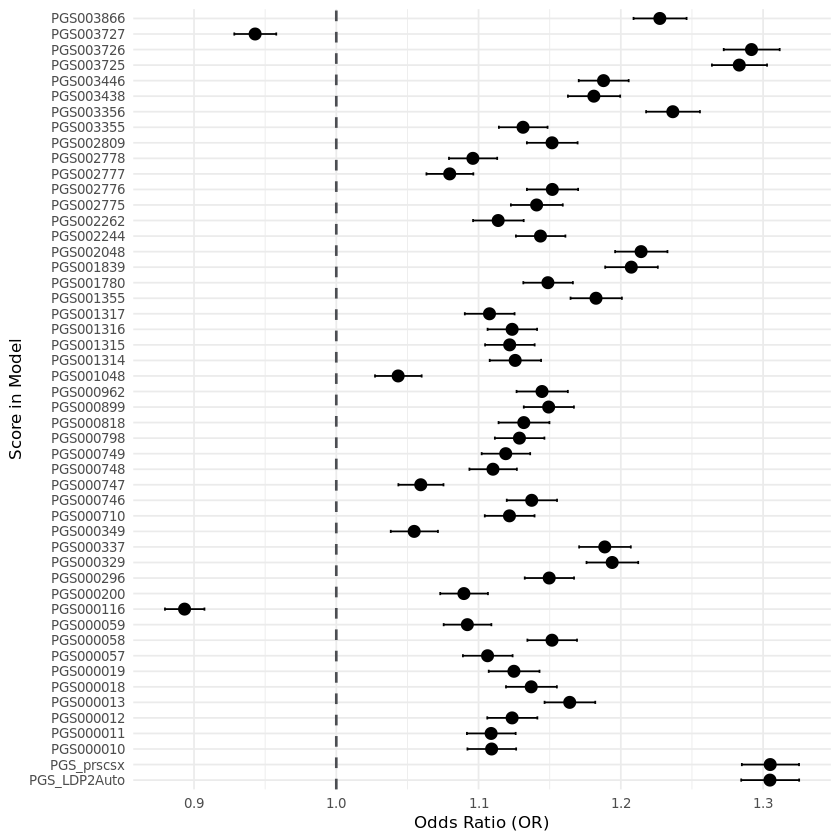

#### **eFigure 12: Forest Plot of Odds Ratios – AOU, PRS Only**

**Forest Plot of Odds Ratios for 50 PRSs in a Model of Prevalent CAD, without covariates, in All of Us.**

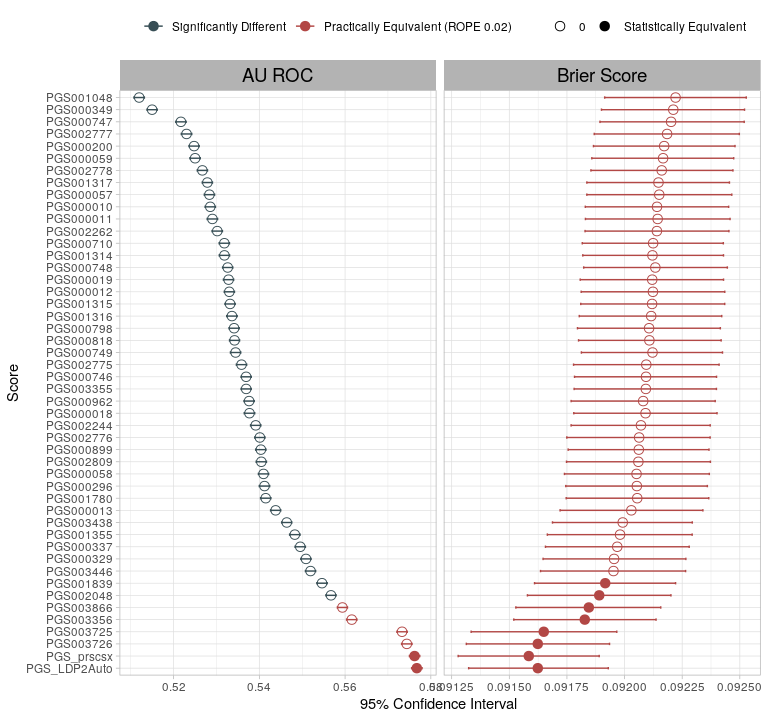

#### **eFigure 13: Distribution of Polygenic Risk Score Brier Scores and AUROCs in AOU – PRS Only**

**Distribution of Polygenic Risk Score Brier Scores and AUROCs in AOU, not including Age and Sex as Covariates.** Estimates and 95% confidence intervals are plotted. Scores in red are practically equivalent using a ROPE of 0.02. Scores with filled in circles have <95% probability of a real positive difference (statistically equivalent). Values are in ST19.

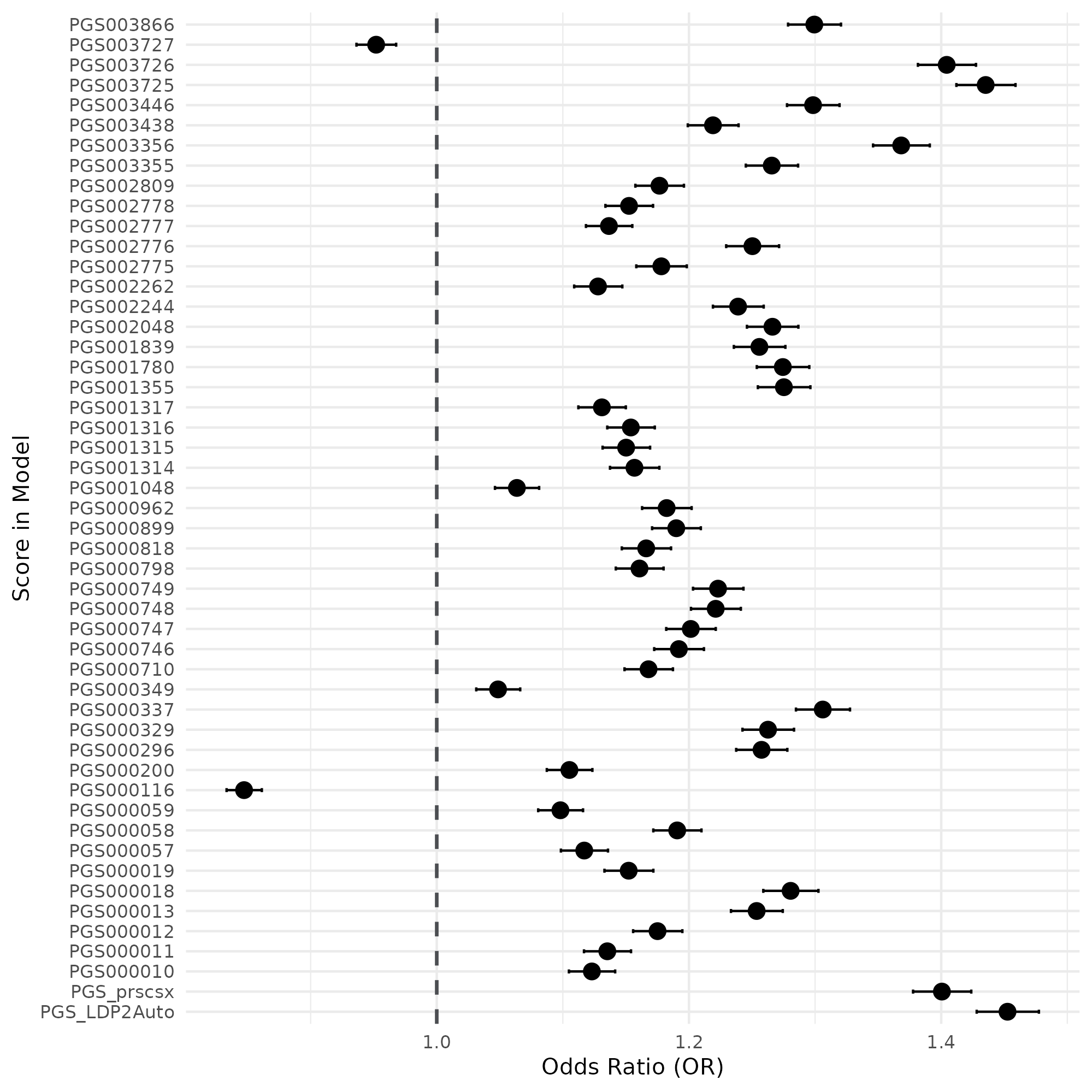

#### **eFigure 14: Forest Plot of Odds Ratios – AOU with PC Covariates**

**Forest Plot of Odds Ratios for 50 PRSs in a Model of Prevalent CAD Including Age, Sex, and First 5 Principal Components, in All of Us.**

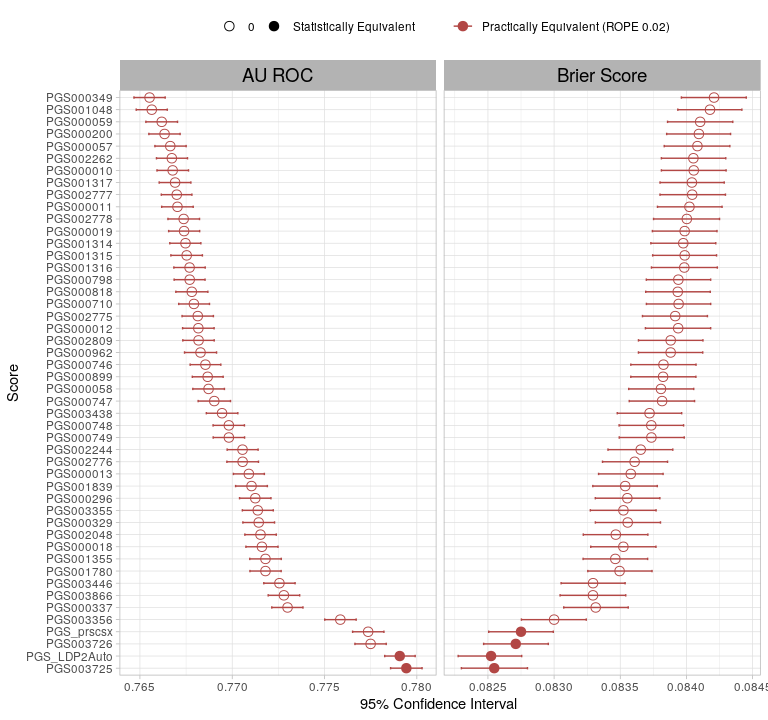

#### **eFigure 15: Distribution of Polygenic Risk Score Brier Scores and AUROCs in AOU with PC Coveriates**

**Distribution of Polygenic Risk Score Brier Scores and AUROCs in AOU, including Age, Sex, and First 5 Principal Components as Covariates.** Estimates and 95% confidence intervals are plotted. Scores in red are practically equivalent using a ROPE of 0.02. Scores with filled in circles have <95% probability of a real positive difference (statistically equivalent). Values are in ST18.

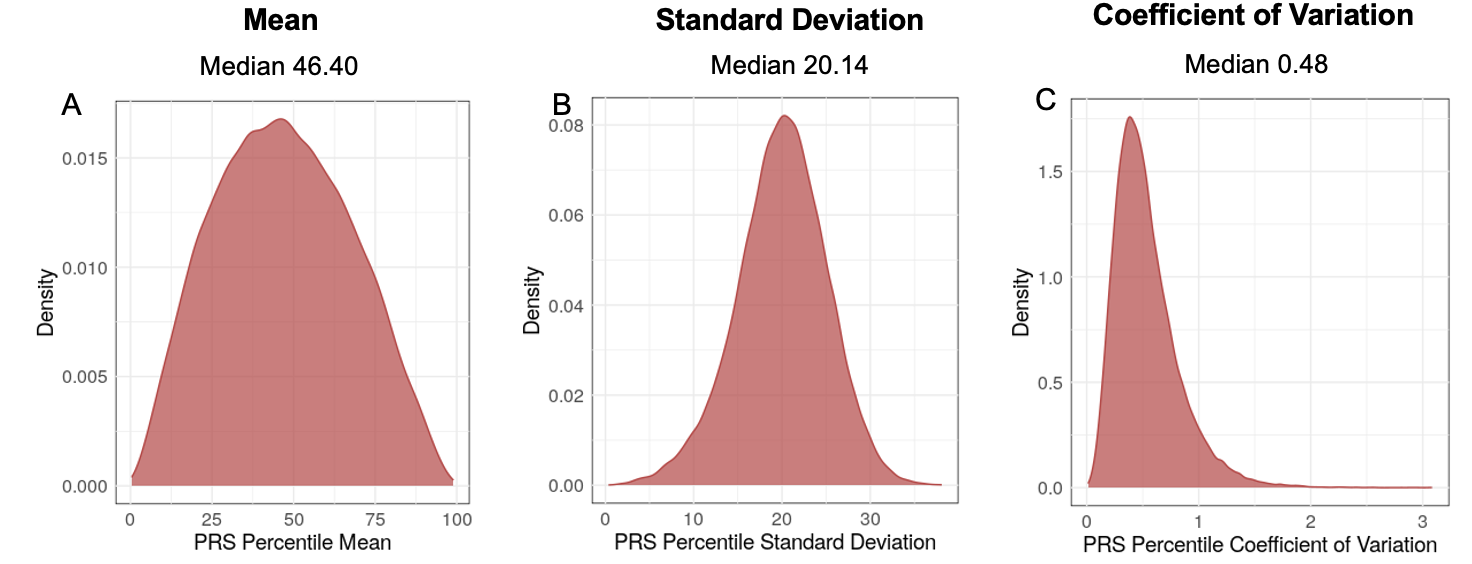

#### **eFigure 16: Within-Person Score Concordance in PMBB.**

Concordance of individual score percentiles across all scores meeting practically equivalent ROPE 0.02 criteria in All of Us (48 scores). **A)** Mean individual risk percentile (median 46.40), **B)** Standard Deviation of the mean individual risk percentile (median 20.14), **C)** Coefficient of Variation (median 0.48).

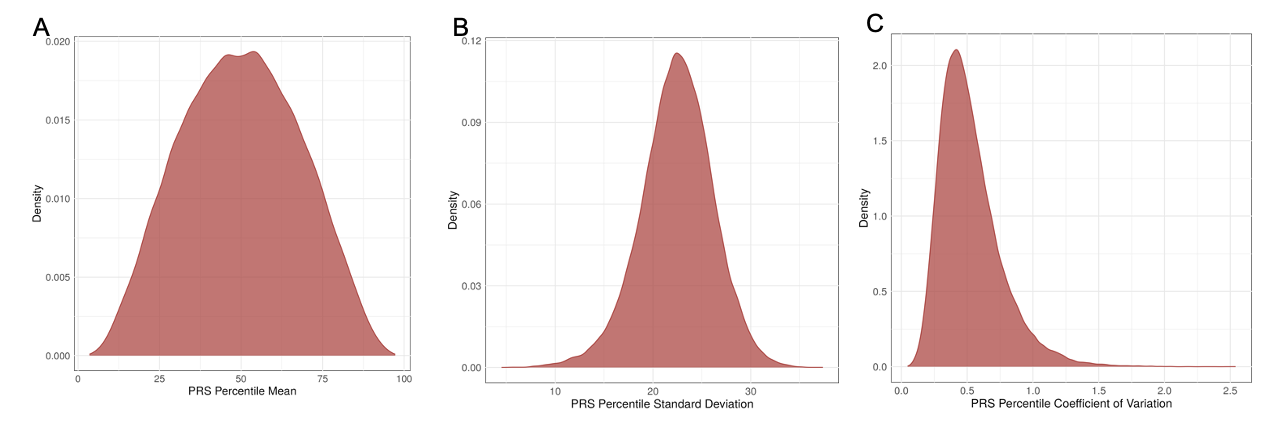

#### **eFigure 17: Within-Person Score Concordance in UCLA**

Concordance of individual score percentiles across all scores meeting practically equivalent ROPE 0.02 criteria in UCLA ATLAS Precision Health Biobank (48 scores). ). **A)** Mean individual risk percentile; median 49.91(49.69, 50.13). **B)** Standard Deviation of the mean individual risk percentile; median 22.55 (22.51, 22.58). **C)** Coefficient of Variation; median 0.4820 (0.4796, 0.4842).

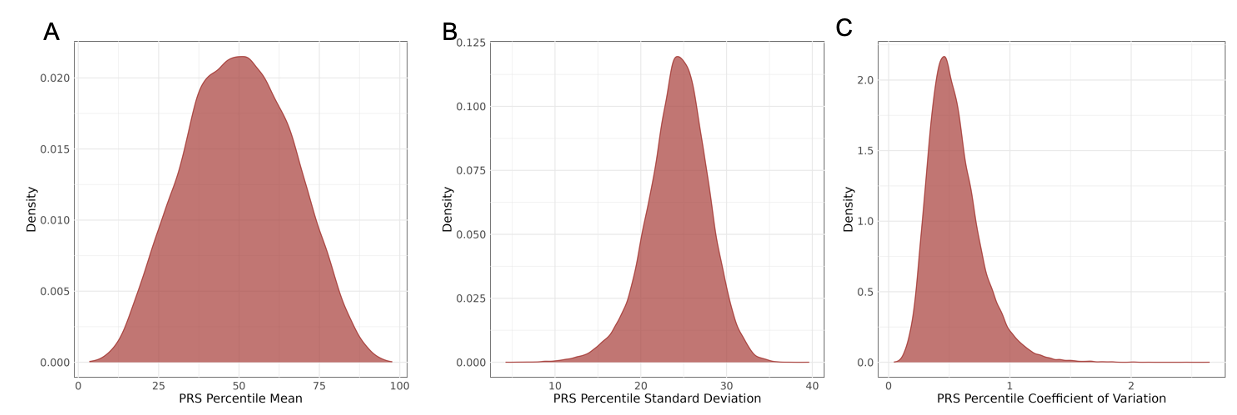

#### **eFigure 18: Within-Person Score Concordance in AOU – AFR**

**Within-Person Score Concordance for Model including Age and Sex as Covariates, in All of Us Individuals most Genetically Similar to a African Reference Population.** Concordance of individual score percentiles across all scores meeting practically equivalent ROPE 0.02 criteria in All of Us (41 scores). **A)** Mean individual risk percentile; median 49.95 (49.71, 50.18). **B)** Standard Deviation of the mean individual risk percentile; median 24.43 (24.39, 24.48). **C)** Coefficient of Variation; median 0.5148 (0.5124, 0.5173).

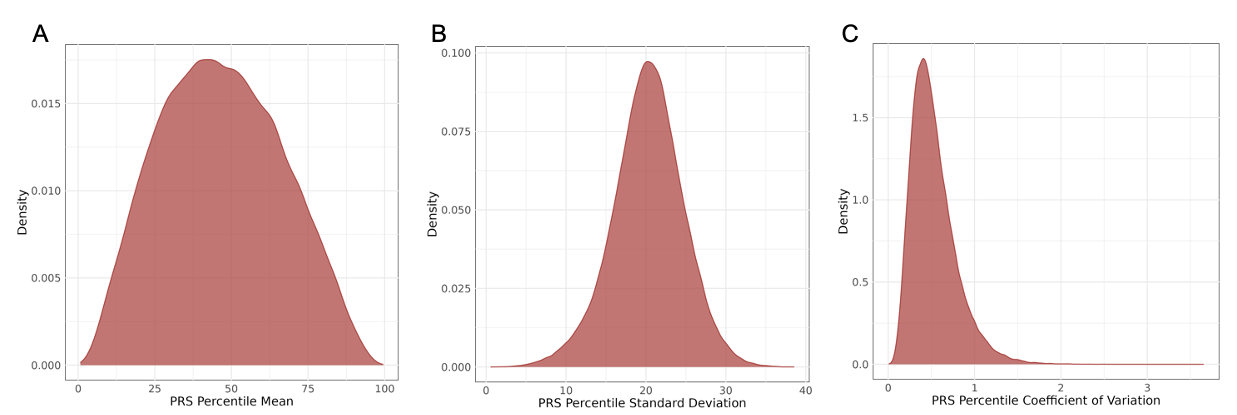

#### **eFigure 19: Within-Person Score Concordance in AOU – EUR**

**Within-Person Score Concordance in AOU for Model including Age and Sex as Covariates, in All of Us Individuals most Genetically Similar to a European Reference Population.** Concordance of individual score percentiles across all scores meeting practically equivalent ROPE 0.02 criteria in All of Us (37 scores). **A)** Mean individual risk percentile; median 46.13 (45.96, 46.29). **B)** Standard Deviation of the mean individual risk percentile; median 20.33 (20.30, 20.36). **C)** Coefficient of Variation; median 0.4808 (0.4789, 0.4826).

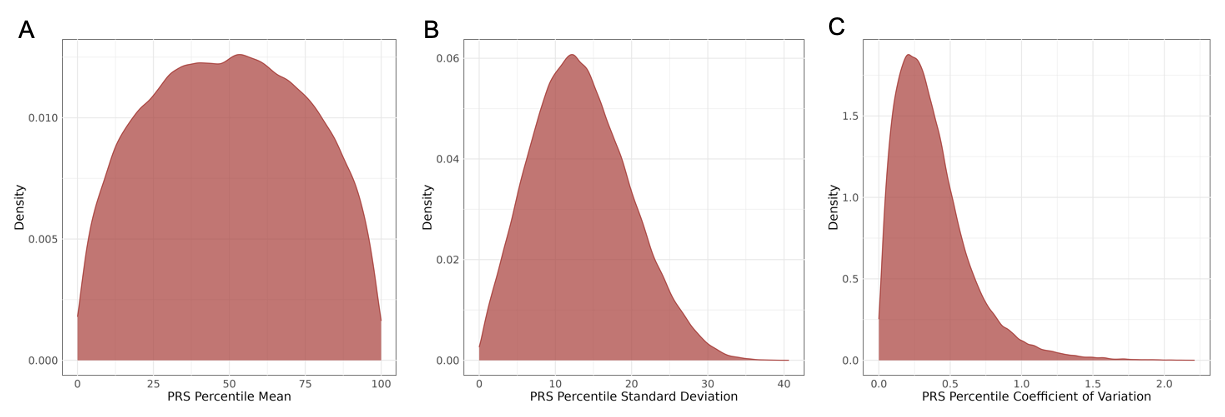

#### **eFigure 20: Within-Person Score Concordance in AOU – PRS Only**

**Within-Person Score Concordance in AOU for Model not including Covariates.** Concordance of individual score percentiles across all scores meeting practically equivalent ROPE 0.02 criteria in All of Us (6 scores). **A)** Mean individual risk percentile; median 49.76 (49.57, 49.96). **B)** Standard Deviation of the mean individual risk percentile; median 13.00 (12.96, 13.04). **C)** Coefficient of Variation; median 0.321 (0.319, 0.322).

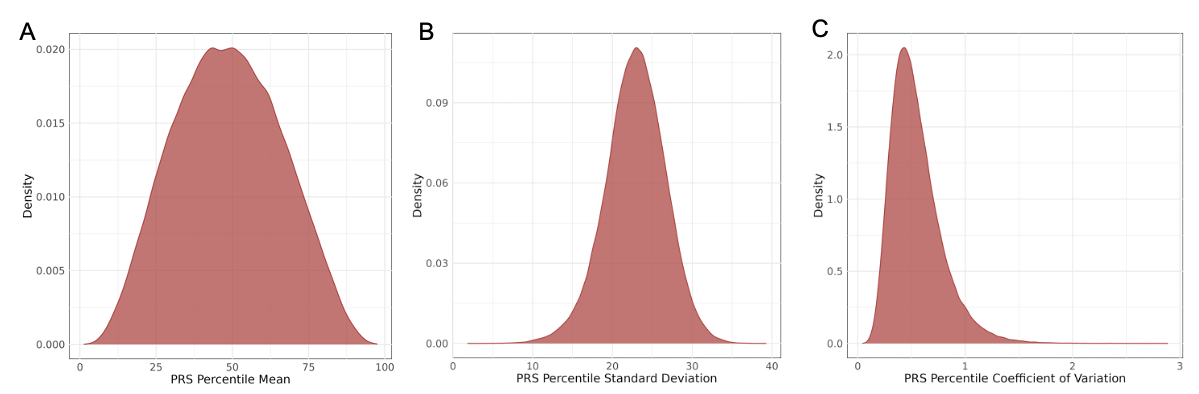

#### **eFigure 21: Within-Person Score Concordance in AOU – Add PC Covariates**

**Within-Person Score Concordance in AOU for Model including Age, Sex, and First 5 Principal Components as Covariates.** Concordance of individual score percentiles across all scores meeting practically equivalent ROPE 0.02 criteria in All of Us (48 scores). **A)** Mean individual risk percentile; median 48.38 (48.26, 48.50). **B)** Standard Deviation of the mean individual risk percentile; median 22.94 (22.92, 22.96). **C)** Coefficient of Variation; median 0.504 (0.503, 0.504).

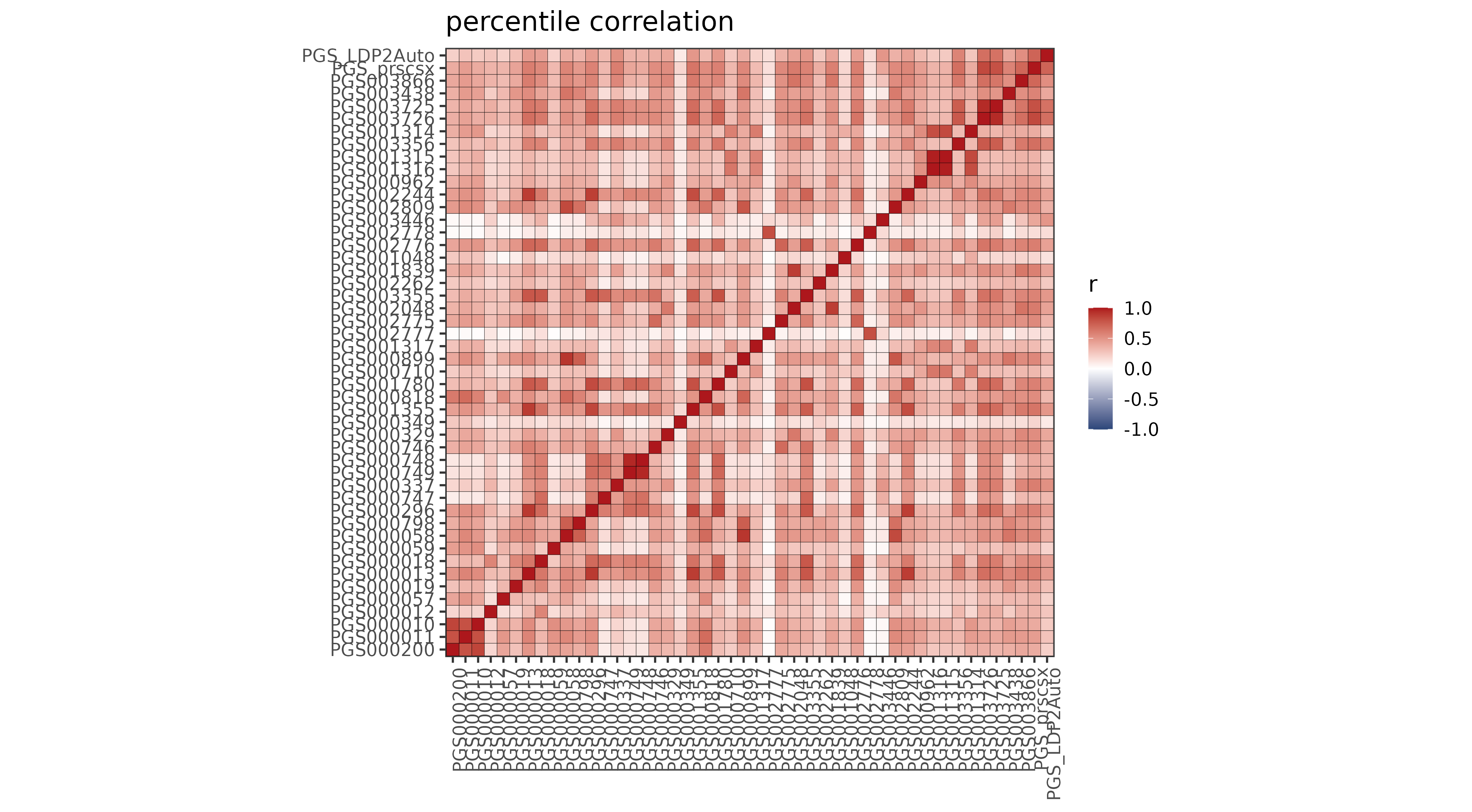

#### **eFigure 22: Pearson r AOU**

**Heatmap of Pearson’s Correlation Coefficient Between Risk Estimates Provided by Pairs of All of Us Scores**

**A**
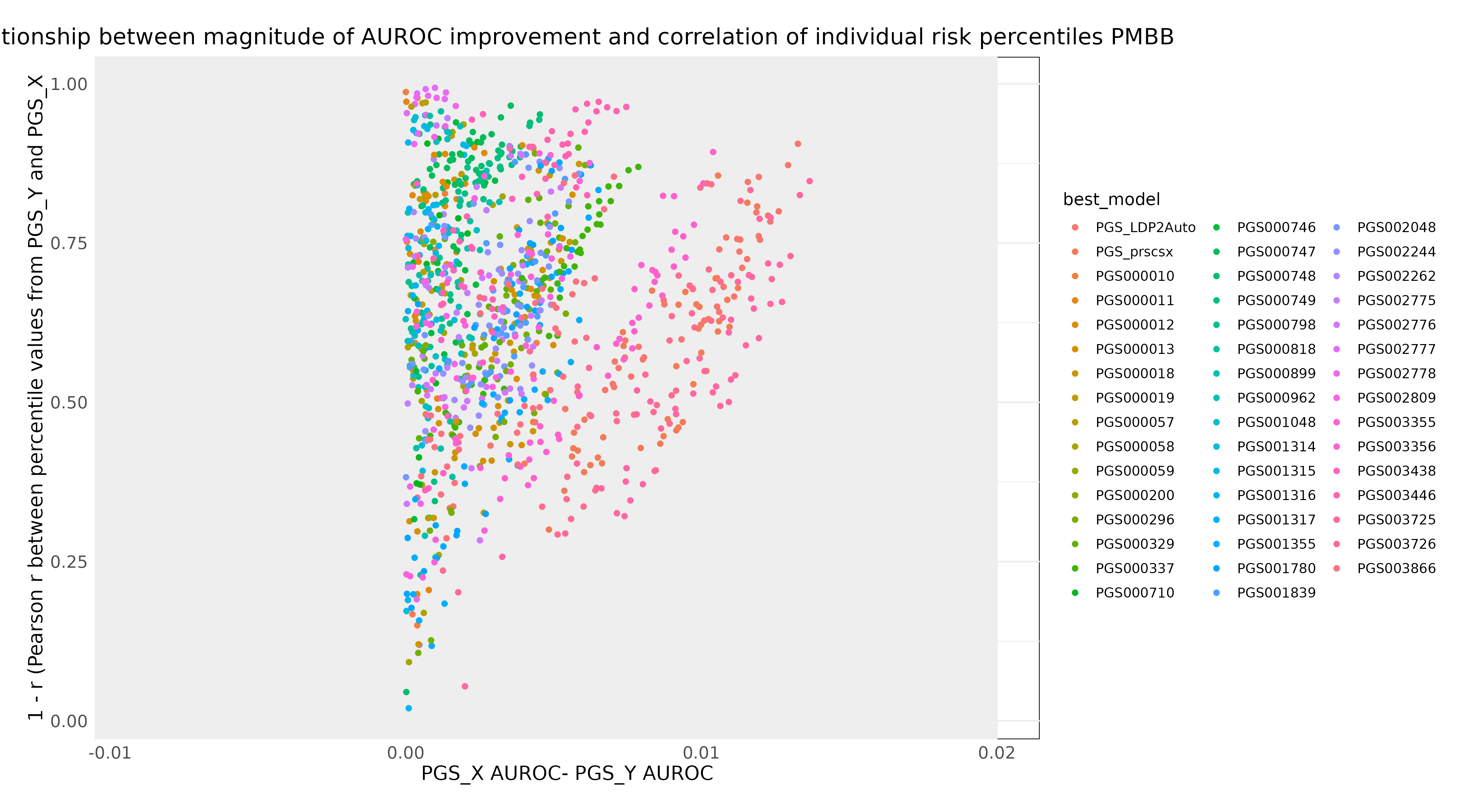

**B**
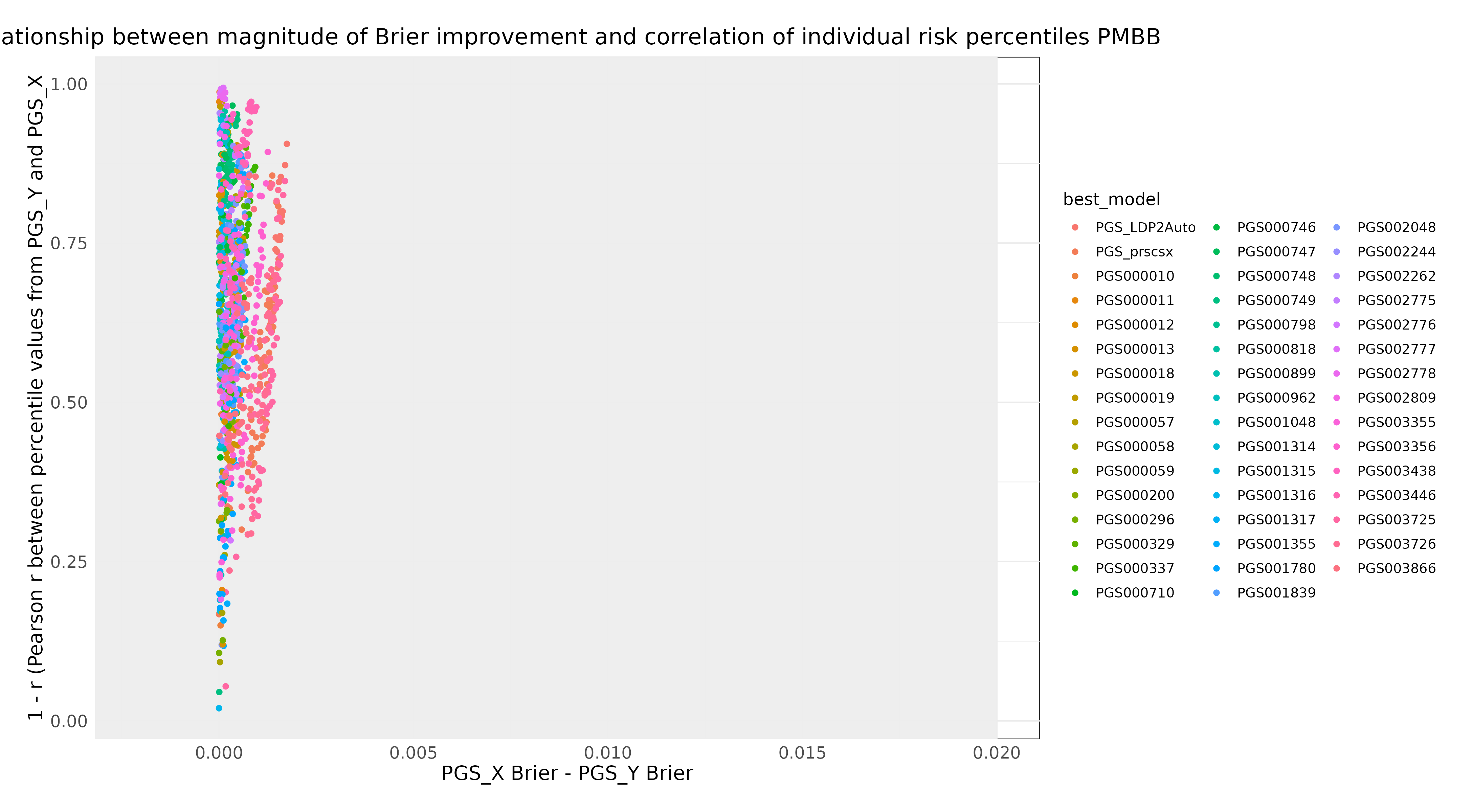

#### **eFigure 23: Score Pair Correlation by Magnitude of Performance Difference - AOU**

**Plot of Score Pair Agreement (measured by Pearson correlation coefficient r) by magnitude of difference between model performance metric.** For all pairwise comparisons, differences in Brier and AUROC were calculated as the difference between the higher and lower score. The ‘better’ score (lowest in the case of Brier, highest in the case of AUROC) is colored by score name. ROPE of 0.02 is plotted with a gray rectangle. **A) AUROC B) Brier Score.**

**Penn Medicine BioBank Banner Author List and Contribution Statements**

PMBB Leadership Team

Daniel J. Rader, M.D., Marylyn D. Ritchie, Ph.D.

Patient Recruitment and Regulatory Oversight

JoEllen Weaver, Nawar Naseer, Ph.D., M.P.H., Giorgio Sirugo, M.D., P.h.D., Afiya Poindexter, Yi-An Ko, Ph.D., Kyle P. Nerz

Lab Operations

JoEllen Weaver, Meghan Livingstone, Fred Vadivieso, Stephanie DerOhannessian, Teo Tran, Julia Stephanowski, Salma Santos, Ned Haubein, P.h.D., Joseph Dunn

Clinical Informatics

Anurag Verma, Ph.D., Colleen Morse Kripke, M.S. DPT, MSA, Marjorie Risman, M.S., Renae Judy, B.S., Colin Wollack, M.S.

Genome Informatics

Anurag Verma Ph.D., Shefali S. Verma, Ph.D., Scott Damrauer, M.D., Yuki Bradford, M.S., Scott Dudek, M.S., Theodore Drivas, M.D., Ph.D.,
